# Geometric approach for non pharmaceutical interventions in epidemiology

**DOI:** 10.1101/2023.05.05.23289577

**Authors:** Laurent Evain, Jean-Jacques Loeb

## Abstract

Various non pharmaceutical interventions have been settled to minimise the burden of the COVID-19 outbreak. We build a framework to analyse the dynamics of non pharmaceutical interventions, to distinguish between mitigations measures leading to objective scientific improvements and mitigations based on both political and scientific considerations. We analyse two possible strategies within this framework. Namely, we consider mitigations driven by the limited resources of the health system and mitigations where a constant set of measures is applied at different moments. We describe the optimal interventions for these scenarios. Our approach involves sir differential systems, it is qualitative and geometrical rather than computational. Along with the analysis of these scenarios, we collect several results that may be useful on their own, in particular on the ground when the variables are not known in real time.

## 1 Introduction

In the pandemic situation, governments have settled policies, based on socio-economic appreciations, field studies, and modelling. The toolbox for the crisis management involved mitigations policies. Numerical simulations suggest that these mitigations measures change the final share *r*_*∞*_ of infected people in the population, sometimes markedly. A possible roadmap for a scientific programme to select non pharmaceutical interventions could be as follows.

1. Discuss the choice of the model. Which models are realistic ?
2. Find the mathematical optimisations for the chosen model.
3. Find the counterpart of the optimisation in real life. Determine possible concrete mitigations that approach the targeted mathematical optimisation.
4. Social analysis. Analyse the public acceptance of the mitigation, the economic impact, and the indirect costs on the population health.

In the present article, we choose a variant of the sir-model. We concentrate on the second item of this roadmap. Our target is to obtain theoretical results, and in particular proved qualitative results. Our results shed light and give an understanding of the the numerical simulations of the spread of a disease. We have a particular interest in qualitative results independent of the input parameters, as these results could be more robust on field with little known parameters. The theoretical background, the constructions, and the mathematical results are exposed in full generality in the supplementary material. The main text is dedicated to a larger audience, it explains, contextualises and illustrates the results with simulations.

Our work started with the article [1]. We were puzzled by some simulations exhibiting final fractions infected depending on the choice of the intensity of preventive measures, via a constant *α* in the next generation matrix. Several remarks were formulated, suggesting qualitative explanations of the phenomena observed on the simulations. For instance, “preventive measures were not imposed from the start and were lifted before the epidemic was over” or “lifting restrictions gradually [can prevent] overshoot “. So it was implicit but clear from the article [1] that an adequate scheduling was required to minimise the burden of the epidemic. However, we could not identify the adequate scheduling in precise mathematical terms, nor could we identify the assumptions required to obtain the qualitative behaviour of the examples. Thus our goal was to clarify and give a general picture of what could mean an “optimal scheduling” of the preventive measures for a pandemic outbreak described with a sir-model.

Our interest for qualitative results was reinforced by contradictory results in the literature with respect to the relevance of an early and strong set of mitigation measures. Whereas overshoot was pointed out as a risk in [1] and implicitly in [8], other other sources [7], [12] advocated for early or strong mitigations to save lives. In contrast to papers where strategies are analysed at fixed dates [7], sometimes to wait for some new drugs or vaccines [8], we are concerned with the very long term. We try to minimise the mortality after an infinitely long time using only non pharmaceutical interventions. In other words, this paper considers non pharmaceutical interventions as an active medical tool to minimise the burden of the epidemic in the long run rather than as a tool to postpone the mortality till some new drug comes on the market.

Here is a summary of our results.

- Even in the absence of medicine to wait for, finite time interventions may be considered to minimise the burden of an epidemic because of the dynamics involved. The situation is analogous to a bike on a sloping road : it is not possible to stop before the low point using a finite time breaking, but breaking is nevertheless useful to avoid moving far beyond the low point due to inertia. In symbols, let *r*_*∞*_ be the ratio of finally infected people and let *R*_0_ be the classical reproduction number. A well scheduled finite time intervention can drive *r*_*∞*_ close to 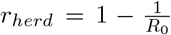, whereas no intervention often leads to *r*_*∞*_ *>> r*_*herd*_. The role of the dynamics is more important when *R*_0_ has a medium value (*R*_0_ ≃2.5) where nearly 30% of the population may avoid the disease thanks to a suitable finite time intervention.
- There is a fundamental qualitative difference between finite time interventions and infinite time interventions. Infinite time interventions can lead to an arbitrarily small ratio *r*_*∞*_ of infected people. In contrast, finite time interventions result in situations where the inequality *r*_*∞*_ *> r*_*herd*_ always holds, so that *r*_*∞*_ close to *r*_*herd*_ is the best possible value.
- Planning is important. Examples show that awkward planning lead to mitigations that are long, costly, with little effect on *r*_*∞*_. The analogy with bikes on a sloping road makes sense again : breaking hard far from the low point hardly has an impact on the inertia and on the distance covered after the low point. In contrast, the analogy with a bike on a flat road is badly suggestive, as it encourages early intensive mitigations with poor results. This leads to a problem of control theory : what are the mitigations which minimise the effort on the population for a fixed result ?
- We build a scientific framework to distinguish between the political level and the scientific level in the decision process. Obviously, the mitigations have a social cost that require a personal subjective appreciation. A rational decision process includes political considerations to aggregate the divergent wishes of the citizen. Nevertheless, some conclusions may be true independently of the subjectivity. Considering two possible choices *A* and *B*, there is a scientific ground to prefer mitigation *A* to mitigation *B* if there are simultaneously fewer infected people and fewer restrictions on the population when *A* is chosen. In contrast, a political trade-off is necessary when a middle ground between infections and constraints has to be found. We keep these notions informal in the main text, but the concepts are rigorously defined in the supplementary material in terms of cost functions and diffeomorphisms. In the following, when we say that a choice *A* is better than a choice *B*, we always refer to the scientific meaning : choice *A* leads both to less restrictions and to less infected people than choice *B*.
- We analyse the scenario where a same constant intervention is applied one or several times. In this context, the two problems of minimising the duration of the constraint for a fixed burden or minimising the burden for a fixed duration are equivalent, and there is no compromise between the duration of the intervention and the number of infections to be found : both are minimised simultaneously. We thus approach the problem with a fixed predefined duration and we determine the adequate planning. This scenario includes for instance the comparison between two strategies, where the first strategy promotes a change every Monday for seven weeks, whereas the second strategy promotes the same change a whole week one month after the starting point of the epidemic. More generally, we compare strategies with a same type of intervention, and the same total duration, and we analyse the optimal planning of the mitigations. We show that in this scenario, splitting the mitigations through several short periods is never optimal. The mitigation minimising the number of finally infected people has always exactly one unique long lasting mitigation. Our model does not support the idea sometimes expressed on the media to plan a strong mitigation as soon as possible. It is quite the opposite. Intensity and timing have to be tuned in a consistent balanced manner : an earlier mitigation must be lighter than an intervention that starts later. Early and strong mitigations are not balanced and yield to poor results. The timing of an optimal planning is understood : it boils down to “as soon as possible” if the herd immunity threshold has been crossed, and around the herd immunity threshold otherwise. Among the possible consistent choices of timing and intensity, the case of a late intensive mitigation, starting when the herd immunity threshold is crossed, has a special interest. The corresponding strategy can be implemented on the ground using measurements in wastewater as in [5]. It may be more easily planned than alternative optimal strategies since estimating *R*_0_ is not necessary, thus bypassing the difficulty of its estimate.
- In a second scenario, we analyse the case where the health system would be saturated in the absence of interventions. Non pharmaceutical interventions are used to maintain the health system below its maximal load. We compare several mitigation strategies with different loads, possibly different from 100%. For instance, the mitigation may start when the health system is filled at 90%. This second scenario is divided in two sub-scenarios :
  − 2a) : The mitigation is shaped so that the health system stays filled at 90% and it is relaxed when the herd immunity threshold is reached. The relaxation occurs when the epidemic naturally decreases.
  − 2b) : This scenario starts like scenario 2a), but the mitigation is relaxed sooner. A rebound of the epidemic occurs and the limit of 90% is exceeded after relaxing, but the total load remains below 100% forever. In other words, the relaxation is launched as soon as returning to normal does not overload the health system in the future despite of a rebound.
  Instead of considering arbitrarily a load of 90% as in the above example, we address the problem of determining the optimal load between 0% and 100% for the health system. What are the optimal loads for scenarios 2a) and 2b) ? First, we show that in scenario 2a), there is no scientific answer. Political trade-offs are unavoidable : a higher load abuts to fewer infected people at the price of more constraints. In scenario 2a), the duration of the mitigation tends quickly to infinity when the considered load goes to zero. Simulations show that the time of mitigation is often very large. It is thus natural to consider scenario 2b) which comes with fewer constraints. We show that, maybe surprisingly, all the strategies considered in the scenario 2b) have the same number of finally infected people, independently of the chosen load. Consequently, a load *A* is preferable than a load *B* if and only if the corresponding strategy leads to fewer constraints on the population for the same result. Small loads are inefficient in scenario 2b). A minimal load is necessary, otherwise the policy is surpassed by other better planned strategies. In the simulations considered, this minimal load of the health system to reach before launching the intervention is large : more than 80% of the maximal load of the health system. This may be viewed as an other incarnation in the context of possibly overloaded health systems of the slogan “A strong and early mitigation is inefficient”. When this minimal load is reached, the question of still enlarging the launching load becomes a political one, it is not a scientific question any more : for the same number of finally infected people, a higher launching load requires an effort for the population which lasts longer, but the maximal effort is lower.
- The above scenarios are built upon a more general analysis which carries several results useful on their own. We give a focus on a function *h* which plays a role similar to energy in physics. In mechanics, a falling object undergoes important damages on the ground if it was thrown with a high kinetic or potential energy. Similarly in our model, if a mitigation is relaxed with a high value of *h*, this will lead to many infected people and fatal cases because of the implied dynamics. Since *h* is an indirect measure of the finally infected people, a mitigation that lowers *h* has a positive impact on the final burden. In contrast, if *h* is only slightly changed by a mitigation, the mitigation hardly has an impact on the final burden : The mitigation is a temporal shift rather than an amelioration, the infections will happen later. A sensible objective for the public policy is thus to lower *h* using interventions of short duration, hence the importance of the derivative 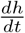. The computation shows that 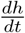 is proportional to the ratio *i*(*t*) of infected people for a fixed intervention. This leads to the very important qualitative result that for a fixed level of constraint, the interventions are more efficient if they occur when many people are infected. The same phenomena has been observed using numerical simulations in [8], however for a different model. This suggests that our qualitative result for the sir model could be extended and could provide an explanation of the numerical observations in other contexts. The variation of the energy function *h* shows that an early intensive intervention acts on mortality similarly to a free loan, with a positive effect on the short term but not on the long term after the loan is repaid. This explains the apparent contradiction between the authors studying the aftermath of the intervention at a fixed date with a positive result [7], whereas [1],[8] have an opposite conclusion with a further time horizon.

### Context and limitations of the findings

Mitigation strategies may be promoted through coercive laws, or they may be exposed to the public as mere recommendations with no obligation. Citizen may have more choice when the intervention occurs (new option for remote work or for a day off for instance) or fewer choices due to restrictions. Our paper is agnostic about the implementations. We consider a model which modifies the coefficients of the sir systems when people modulate their interactions, but we make no assumption on the social tools used to modify these coefficients.

Several questions have not been considered.

We do not discuss the economical or global health impact nor the social acceptance in this paper.

Field testing, studies with animal models and experiments to support the numerical and theoretical results would be welcome. Insect pathogens have been used to test equations because of their tractability [2, 4]. Minimising the spread of the epidemic in plants while minimising the intervention is a natural question, and we have not explored how our results could be enlightening for agriculture or other epidemiological contexts.

These questions are out of the scope of the article, and they belong to a field of work where we have no expertise. However, we believe that these are important questions to be discussed by qualified researchers in these fields.

We discuss now the choice of the model. Some modellings rely on simple models, whereas other modellings require many interacting parameters. Both have their pros and cons, depending on the objectives, quantitative or qualitative, and on the quality of the data. As a rule of thumb, simple models with few variables allow qualitative explanations, they have a lower sensitivity to the parameters. When high quality data and model is available, complex models with more variables lead to more precise predictions. Qualitative interpretation of the changes implied by the modifications of the input constants is difficult for complex models. Both approaches are complementary rather than opposite.

Since our goal was to identify the qualitative phenomena that drive and circumscribe the computations, a variant of the simple and robust sir-model was a sound choice. In the variant considered, a coefficient that was constant in the original sir model varies with the mitigation policies implemented. Although the sir model is suitable for qualitative analysis, our modelling carries the simplifications and limitations attached to this model : people are infected only once, deaths are not considered, the population is supposed to be geographically homogeneous, all individuals are equally susceptible, viruses undergo no mutations, to cite a few limitations.

There are slight variations of the sir-model for which we can carry our results or follow an approach along the same lines (remark 19). However, there are other models to experiment to approach reality, and they may be quite different. For instance, “on the experimental side, Dwyer et al. (1997) measured nonlinear relationships between transmission and densities of susceptible hosts, implying that the bilinear term in the classical susceptible-infected-recovered (SIR) model may not be appropriate. “ [3, 2]. Many variations are possible, and probably many are necessary depending on the problem under consideration. Closed formulas for a differential system are an exception. Most variations from the SIR-system will lead to models which are not computable with closed formulas. While a large part of our analysis is geometrical and clarifies the involved phenomenons, other arguments still depend on the closed formulas of the sir system and will not apply to non computable models. Generalisations based on a better understanding of the geometrical architecture could be explored, to prove our results for a larger class of models.

Many parameters are not constant, their value evolves with time. For instance, we do not know how many people will be vaccinated, how often, and the efficiency of the vaccines on the variants to come. In this rapidly changing environment, we hope that our qualitative results may be useful, in particular those independent of the numerical data in input.

## 2 Main text

### Mitigation as an active tool

In a pandemic context, mitigation policies are usually understood as a tool to give deciders and physicians time for dealing with the problem. Delaying the epidemic gives time for researchers to find new remedies and gives time to setup the logistics to vaccinate people. Our approach in this paper is different. We consider mitigations as a tool to minimise the number of finally infected people, even in the absence of change in the remedies. The goal of this article is to develop this point of view of mitigation strategies as an active tool for driving the epidemic. In this section, we exhibit simulations to expose the problematic involved.

A mitigation that improves the final situation without any new drug is illustrated in the next figure. The red drawing is the trajectory of the pandemic without mitigation. The green drawing is the same pandemic, where a mitigation is applied from week 22 to week 26. During the four weeks of mitigation, the reproduction number *R*_0_ has been reduced from 2.4 in the absence of mitigation to *R*_1_ = 0.4. Thanks to the mitigation, the share *r*_*∞*_ of finally infected people dropped from 0.88 to 0.82.

The evolution of the pandemic is shown in the (*s, r*)-plane, more precisely in the triangle *s >* 0, *r* ≥0, *s*+*r*≤ 1. Here *s* and *r* are the share of susceptible people and the share of removed people in the population, with the standard notations of the sir model. The share *i* of infected people is implicit since *i* = 1 −*r* −*s*. The point *M* (*t*) = (*s*(*t*), *r*(*t*)) that represents the epidemic at time *t* starts at *t* = 0 with *M* (0) at the bottom right (*M* (0) (1, 0)). As *t* increases, *M* (*t*) moves to the left (*s* decreases) and the limit point *M*_*∞*_ = (*s*_*∞*_, *r*_*∞*_) is on the diagonal *r* + *s* = 1 (*i*_*∞*_ = 0). Note that by construction *r*_*∞*_ is the share of people that have been infected at some time *t* during the epidemic, with 0 *< t < ∞*. The limit point *M*_*∞*_ is lower for the green curve than for the red curve. This means means that the mitigation has been efficient, it has lowered *r*_*∞*_.

What is a good mitigation strategy ? The problem can be thought in analogy with the braking of a bike on a slope. All braking strategies are not equivalent : a rider does not apply a constant braking force on downslopes. There are moments where braking is useless, and other moments where braking is necessary to take the turn. Technically, this is a question of control theory. The riders on the Tour de France unconsciously apply some control theory to find the optimal timing and intensity for the braking.

The same phenomenon appears in an epidemiological context. A mitigation measure is the analogue of a braking action to slow down the epidemic. Since mitigations limit the possibilities for citizen, one wants to minimise the duration and intensity of these mitigations for the same braking performance, or to minimise the number of infected people for a fixed level of braking effort.

A naïve approach would suppose that control theory is straightforward, that no planning is required, and that the value of *r*_*∞*_ depends only on how strict and how long the mitigations are. This is not the case : adequate scheduling is important. A lighter and shorter mitigation may outperform a harsher mitigation thanks to a better scheduling, as illustrated with the following example. The mitigations for the green trajectory are shorter-lived than those for the red trajectory (4 weeks vs 6 weeks), are less intensive (reproduction during the mitigation R1=0.8 vs R1=0.7), yet the share *r*_*∞*_ of finally infected people is lower for the green curve. A public policy should recommend the green trajectory over the red trajectory.

## 3 Orders of magnitude

What is the difference between a perfect mitigation and no mitigation ? How many saved lives and how many people may avoid infection using an efficient mitigation ? We give some estimates in this section.

In a free environment without mitigation, the share *r*_*∞,free*_ of finally infected people in a sir model is the unique positive solution of the equation

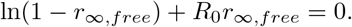

(Theorem 16). With an optimal mitigation, the ratio of finally infected people is about

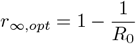

(Theorem 12). The share Δ_*r*_ of the population avoiding an infection with an optimal mitigation is thus

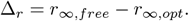

The share Δ_*deaths*_ of lives saved is

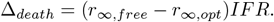

where IFR denotes the infection fatality rate.

Some estimates of these quantities are given in the table of figure 3 for different values of *R*_0_ and *IFR*. When *R*_0_ is slightly above 2 : up to 30% of the general population avoids an infection with an optimal mitigation. This high figure means that, from a mathematical point of view, mitigation as a tool to reduce the burden of an epidemic makes sense. There is no guarantee however that this strategy is possible in real life.

**Figure 1:**
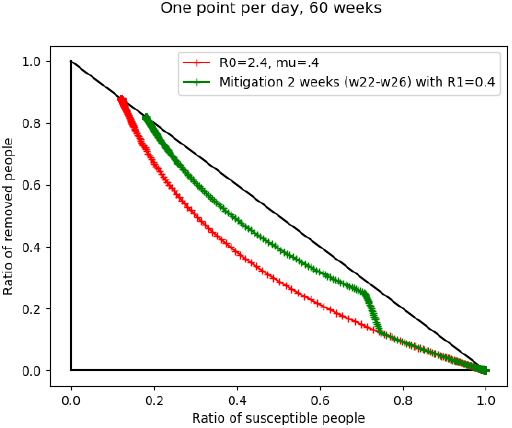
A simple mitigation

**Figure 2:**
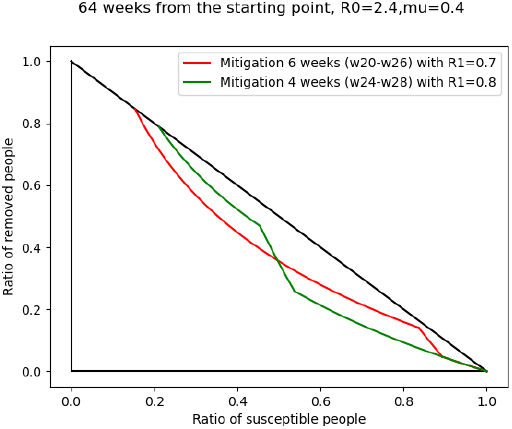
Two mitigations with different plannings.

**Figure 3:**
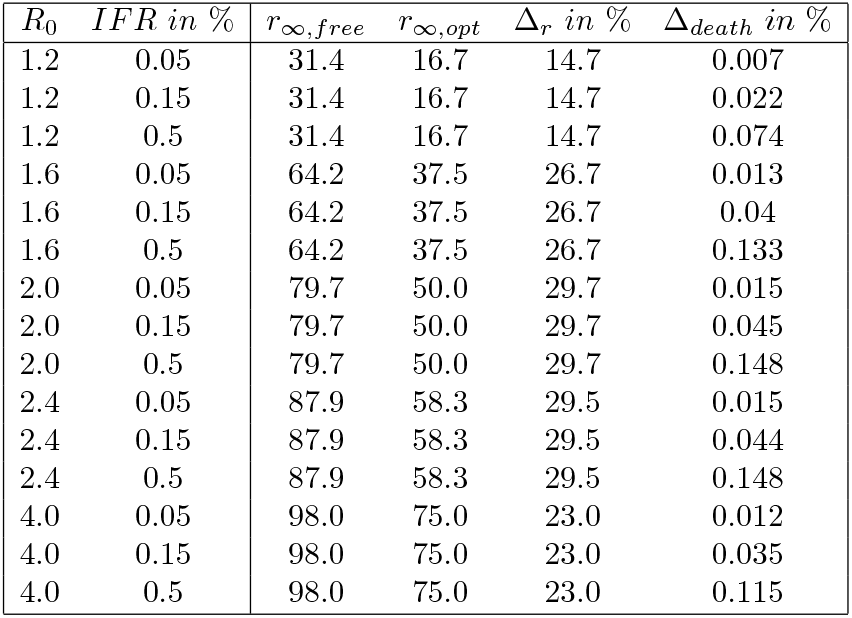
Orders of magnitude

We remarked quite surprisingly that the importance of mitigation increases for medium *R*_0_. The natural guess that mitigation should play a more important role for *R*_0_ large is wrong : for large *R*_0_, the difference Δ_*r*_ = (*r*_*∞,free*_ − *r*_*∞,opt*_) is small because 0 *< r*_*∞,opt*_ *< r*_*∞,free*_ *<* 1 and 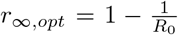 is close to 1. This phenomenon is illustrated in figure 4 showing Δ_*r*_ as a function of *R*_0_.

**Figure 4:**
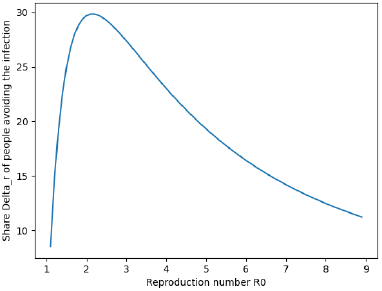
Δ_*r*_ as a function of *R*_0_

Note that the figures in table 3 for Δ_*r*_ are only upper-bounds for the objectives of the public policies. People often react naturally when a brother or a friend is ill. In a growing epidemic, the population limits its interactions by itself. For instance, it was remarked in [10] that “most of the decline in mobility in [the] sample happened before the introduction of lockdowns. Failing to account for voluntary changes in behaviour leads to substantially over-estimated effects of non pharmaceutical interventions ”. We call Δ_*nat*_ the share of the population avoiding an infection due to this reaction of the public. The share Δ_*public policy*_ of the population protected against infection by the public policies is in addition to Δ_*nat*_. The population that avoids an infection thanks to mitigation is Δ_*nat*_ + Δ_*public policy*_. If the combined effect of natural reaction and public policies is optimal, Δ_*nat*_ + Δ_*public policy*_ = Δ_*r*_. Without the optimality hypothesis, Δ_*public policy*_ ≤ Δ_*r*_ −Δ_*nat*_. In our views, the role of the political institutions is to coordinate and amplify if necessary the natural movement of the public to maximise Δ_*public policy*_, in accordance to the history and social context of the country. Planning and scheduling the information to the public is an ingredient of this maximisation, as media campaigns can increase the level of compliance to safe attitudes at the appropriate time. Prophylactic measures are dependent on the mode of transmission of the disease. Promoting the correct prophylactic attitudes at the key moments could also be an objective of the public policy.

### Studying scenarios

Our next goal is to give a qualitative analysis of the involved phenomena during the mitigations. To this aim, we tried to go beyond numerical simulations, because their qualitative interpretation is often difficult, and because extracting a general behaviour from examples depending on the input data is in our opinion a slippery methodology. Rather, we consider two scenarios, and we prove qualitative results that apply independently of the numerical data in input. The first scenario has fixed mitigations. The second scenario considers situations where the health system is overwhelmed in the absence of mitigation.

In the first scenario, we consider a fixed action : for instance a larger part of the population works remotely. This leads to a mitigation with reproduction number *R*_1_ which is lower than the initial reproduction number *R*_0_ *>* 1. Both cases *R*_1_ *>* 1 and *R*_1_ *<* 1 make sense. We fix a total duration *d* for the mitigation measure. This whole mitigation is split in *k* + 1 shorter uninterrupted mitigations of duration *d*_0_, …, *d*_*k*_. The total duration of the mitigation is the sum of the duration of the uninterrupted mitigations, hence ∑*d*_*i*_ = *d*. In this context, the question is : what is the optimal value for the number *k* and when should these *k* + 1 mitigations occur ? Our answer is that the optimal strategy satisfies *k* = 0. The optimal strategy is an uninterrupted mitigation, which is not split in several shorter mitigations. This is a general fact independent of the values of *R*_0_ and *R*_1_. It is illustrated in the left part of figure 5 : a 60 days mitigation lowers the reproduction from *R*_0_ = 2.6 to *R*_1_ = 0.4 (*μ* = 0.4). This 60 days mitigation is not split (blue curve, partially hidden by the orange curve), split in two shorter mitigations of 30 days (orange curve), or five shorter mitigations of 12 days (green curve). The pause between two successive mitigations is three times the duration of the mitigations, namely 90 days (orange curve) and 36 days (green curve). For each of these three scenarios, the start time for the first mitigation has been chosen to give the smallest possible *r*_*∞*_. If the start time of the first mitigation is changed, the value of *r*_*∞*_ depending on the start time (in weeks) is given in the right part of the figure for each scenario. For instance, for two mitigations of 30 days distant of 90 days, the orange curve on the right part of figure 5 says that in our simulations, the optimal start time for the first mitigation is a little more than 8 weeks after the first few imported cases, and yields to *r*_*∞*_ between 0.775 and 0.8. The corresponding epidemic is drawn on the left side.

**Figure 5:**
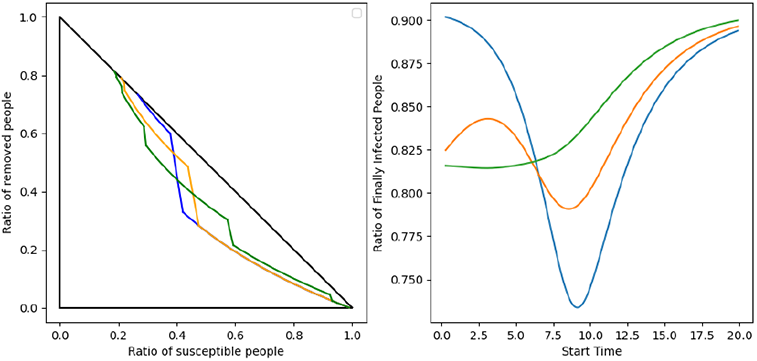
Splitting mitigations

Moreover, in the unsplit case *k* = 0, we have an estimate for the optimal moment for the intervention. Optimality requires that the equality 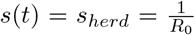 occurs at a moment *t* during the mitigation. This is also a general result independent of the numerical values of *R*_0_ and *R*_1_. If the mitigation ends at a time *t* with *s*(*t*) *< s*_*herd*_, it is too early. If the mitigation starts with *s*(*t*) *> s*_*herd*_, it is too late (Theorem 28). In the example of the figure, with *R*_0_ = 2.6, we have *s*_*herd*_ = 0.38. The optimal strategy illustrated by the blue curve of the figure has numerical results consistent with the general theorem : the vertical line *s* = *s*_*herd*_ = 0.38 is crossed during the mitigation period, represented by the most vertical part of the left blue curve.

Using the analogy with a bike on a slope, and considering that the low point on the road is the analogue of the herd ratio, our theorem says that the optimal breaking occurs in one step, and we should be breaking in a zone which encompasses the bottom of the slope. Ending the breaking before the lowest point of the valley would be inefficient, starting the breaking after the low point would be inefficient too.

In the scenario considered so far, the constraint was the same for all strategies (same restrictions, same total duration for the mitigation). In the next scenario, it will be necessary to compare heterogeneous constraints, which are not constant in time nor have the same duration. The comparison is easy in some cases. For instance, it is less constraining to have a soft mitigation lasting two days than a harsh mitigation lasting five days. For some other cases, the comparison is not possible : There is no natural choice between a long soft constraint, and a short harsh constraint. Finally, there are comparisons which are possible but may require a moment to reflect. As an example, a strategy *S*_1_ imposing a partial set of constraints for 2 days and a total set of constraints for 3 days is less constrained than an alternative strategy *S*_2_ imposing the same partial mitigation for 1 day, and the same total mitigation for 5 days (reason: *S*_2_ is obtained from *S*_1_ by replacing one day of partial constraints with two days of complete constraints). This approach to order the constraints on the above examples can be formalised and written rigorously. In the appendix, we formalise and extend these ideas to compare the constraints of two different strategies *S*_1_ and *S*_2_ to the case of mitigations whose constraints vary continuously with time. To keep things simple yet intuitive, there are fewer constraints for the strategy *S*_1_ than for the strategy *S*_2_ if every person prefers *S*_1_ than *S*_2_, whatever her personal cost function. This occurs in particular when *S*_2_ is obtained from *S*_1_ by replacing mitigations of duration *d* with harsher mitigations of duration *d′ > d*.

In the second scenario, we consider a risk of overwhelmed health systems. We fix an upper bound *i*_*trig*_ for the maximal ratio of infected people. For instance, *i*_*trig*_ = 0.1 means that when 10% of people are infected simultaneously, a mitigation is triggered to prevent the increase of the number of hospitalised patients. The level of mitigation is then settled so that the ratio of infected people stays exactly at *i* = *i*_*trig*_ = 0.1. Equivalently, there are as many people getting sick as people being cured by unit of time during the mitigation period. The process is illustrated in the next figure. The epidemic starts with very few infected persons, and the initial propagation is drawn in red. Then, the level of infection which triggers the mitigation measures is reached and the mitigation period with constant *i* corresponds to the blue segment. After some time (13 weeks in the example of the figure, *μ* = 0.33), the level of infected people naturally decreases. The mitigations are relaxed as they are not necessary any more (in orange on the figure). A similar strategy can be set up with an other value for *i*_*trig*_. The question in this context is the determination of the optimal *i*_*trig*_ ?

We show that there is no scientific answer to this question. A political trade-off is necessary, as minimising constraints and minimising the ratio *r*_*∞*_ of finally infected people are opposite objectives. A small *i*_*trig*_ corresponds to a smaller *r*_*∞*_ at the price of more constraints (Theorem 40). Figure 7 illustrates this fact with two mitigation levels *i*_*trig*_ = 5% and *i*_*trig*_ = 15%. The red curve represents an epidemic without mitigation. The two possible mitigations are drawn in blue. The ratio *r*_*∞*_ is smaller for *i*_*trig*_ = 5%, but the mitigation lasts far longer than for *i*_*trig*_ = 15% (35 weeks vs 8 weeks with *R*_0_ = 3, *μ* = 0.33). Moreover, the initial reproduction number of the mitigation (defined as the reproduction number when the mitigation has just started) is smaller for *i*_*trig*_ = 5% (1.08 vs 1.32). This means that harsher and longer constraints are necessary for a small *i*_*trig*_ value.

**Figure 6:**
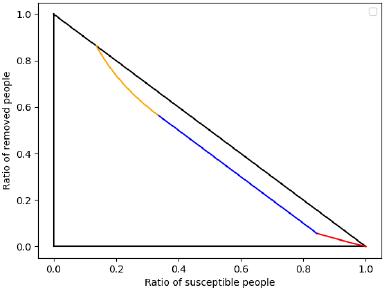
Fixing the maximum level of infected people

**Figure 7:**
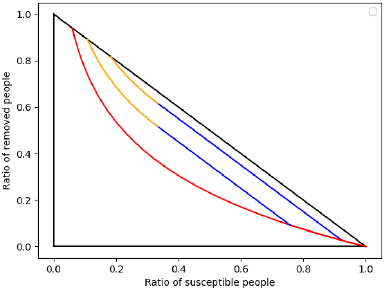
Comparing two different values of *i*_*trig*_.

As *i*_*trig*_ tends to 0, the constraint duration tends rapidly to *∞*. This is illustrated in figure 8 where the duration of the mitigation in weeks is plotted in red, and the initial reproduction number defined above is plotted in green (scaled by a factor 100). Both are plotted as functions of *i*_*trig*_.

**Figure 8:**
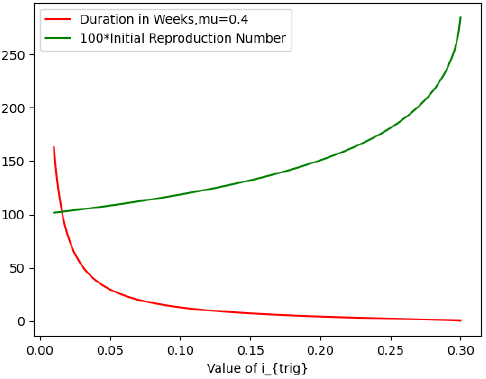
Duration and intensity of mitigation as functions of *i*_*trig*_ when *R*_0_ = 3.

To minimise this long constraint when *i*_*trig*_ is small, we consider a variant of this scenario. For this variant, when the ratio *i* of infected people reaches *i* = *i*_*trig*_, a mitigation is set up as above to preserve the health system. But the mitigation is relaxed sooner in comparison to the previous scenario : as soon as it is possible to stop the mitigation without overwhelming the health system in the future, the mitigation is relaxed. For instance, suppose that the health system is totally full when *i* = *i*_*hosp*_ = 0.15. A mitigation is triggered when *i* = *i*_*trig*_ = 0.05, and it is maintained for a moment so that *i*(*t*) stays blocked at the constant value *i* = 0.05. When the mitigation is relaxed, the ratio *i* of infected people increases again from *i* = 0.05, but it never exceeds *i*_*hosp*_ = 0.15. In other words, a rebound of the epidemic occurs but the health system remains not full and viable after the mitigation is over. This strategy 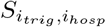 is illustrated in figure 9. It depends on the two constants *i*_*trig*_ and *i*_*hosp*_. The constant *i*_*hosp*_ depends on the health system of the country and is not changeable in the short term. In contrast, different values of *i*_*trig*_ are possible and lead to different public policies.

**Figure 9:**
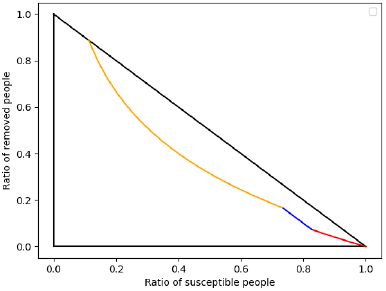
A mitigation is settled, then relaxed with a possible rebound.

The duration of the mitigation in the scenario 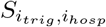 is shown as a function of *i*_*trig*_ in Figure 10. We see that the duration in weeks (represented by the blue curve) is shorter when a rebound is allowed, in comparison to the previous scenario without rebounds (orange curve). The difference is significant, thus the objective of lowering the time of mitigation by allowing a rebound is achieved.

**Figure 10:**
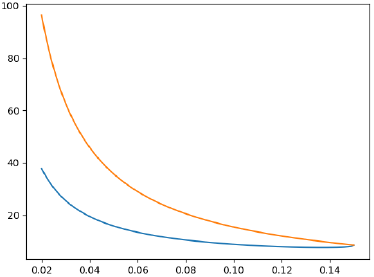
Duration of the mitigation in weeks as a function of *i*_*trig*_ for *i*_*hosp*_ = 0.15, *R*_0_ = 3.

Besides a shorter mitigation time, there are several differences that make the scenario with rebound quite different from the scenario without rebound.

First, when a final rebound is allowed, the duration is not a decreasing function of *i*_*trig*_ any more, as illustrated by a zoom on the previous blue curve (Figure 11). The minimal duration of around 7.6 weeks for the mitigation is obtained for *i*_*trig*_ = *i*_*min*_ := 0.136.

**Figure 11:**
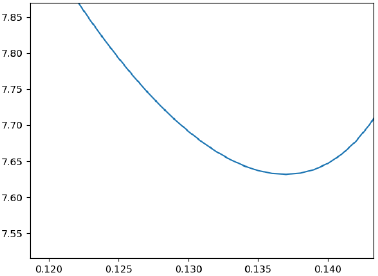
Duration of the mitigation in weeks as a function of *i*_*trig*_ for *i*_*hosp*_ = 0.15, *R*_0_ = 3.

Second, the variation of *r*_*∞*_ is different too. In the scenario without rebound, an early intervention lowered the ratio *r*_*∞*_ at the price of a longer and harsher mitigation. In the scenario with rebound, a harsher or longer mitigation is not rewarded by a smaller *r*_*∞*_. All strategies 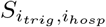 have the same ratio *r*_*∞*_ of finally infected people independently of *i*_*trig*_ for a fixed hospital capacity *i*_*hosp*_. In other words, the level *i*_*trig*_ that triggers mitigations does not influence how many people will be finally ill or dead (Theorem 44 in the appendix). This surprising phenomenon is illustrated in figure 12 with *i*_*hosp*_ = 0.15 and two different values of *i*_*trig*_. The mitigations in blue are triggered when *i*_*trig*_ = 0.05 and *i*_*trig*_ = 0.10 respectively. They are relaxed as soon as *i*_*hosp*_ is never exceeded. The value of *r*_*∞*_ which is common to the two mitigations is the *r*-coordinate of the point at the end of the yellow curve.

**Figure 12:**
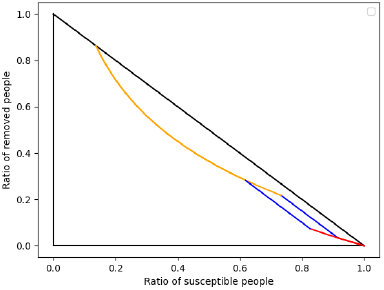
Mitigations with *i*_*hosp*_ = 0.15, *i*_*trig*_ = 0.05 or 0.10.

As a consequence, the mitigations with *i*_*trig*_ *< i*_*min*_ must be rejected. Indeed, they are longer and harsher than the mitigation with *i*_*trig*_ = *i*_*min*_ and the supplementary constraint is not compensated by an amelioration of *r*_*∞*_.

We have excluded the cases *i*_*trig*_ ∈]0, *i*_*min*_[. Let us now consider the remaining range *i*_*trig*_ ≥ *i*_*min*_. In this range, all values of *i*_*trig*_ may be considered and lead to non comparable mitigations. More precisely, if mitigations are triggered above the minimal load *i*_*min*_, then a higher load *i*_*trig*_ gives a longer but less intensive mitigation. Thus a political trade-off between intensity and duration of the mitigation is required to make the choice. Some people may prefer a short intensive mitigation (*i*_*trig*_ close to *i*_*min*_) while other people may prefer a long cool mitigation (*i*_*trig*_ close to *i*_*hosp*_).

The remarks formulated on the example are illustrations of general qualitative results proved in Theorem 44. The theorem can be summarised as follows. In the variant with a rebound allowed, the choice of the level *i*_*trig*_ which triggers the mitigation has no impact on the share *r*_*∞*_ of finally people infected. The choice of *i*_*trig*_ impacts only the subjective human or economical cost of the mitigation, but the direct burden of the disease remains unchanged. There exists a minimal load *i*_*min*_ characterised by the following properties. If *i*_*trig*_ *< i*_*min*_, the strategy is to be rejected as the mitigation is unnecessarily long and harsh for the same result. All the choices with *i*_*trig*_ ∈ [*i*_*min*_, *i*_*hosp*_] are possible and correspond to different trade-offs between length and intensity : *i*_*trig*_ closer to *i*_*min*_ corresponds to a shorter and harsher mitigation. In the example with *i*_*hosp*_ = 0.15, we have *i*_*min*_ = 0.136. Since *i*_*min*_ is close to *i*_*hosp*_, this means that the mitigation must be triggered when the health system is nearly full. Other simulations also give *i*_*min*_ close to *i*_*hosp*_. These simulations express the idea that, for the scenario with rebound within a sir-system, it is a wrong idea to anticipate much and to launch mitigation measures far before the saturation of the health system.

### Temporary versus definitive mitigations, and temporal shifts versus improvements

The idea “the sooner the restrictions, the better” is often implicit or explicit in the debate. For instance in [6], several epidemiologists called for early mitigation measures. It is not supported by the above simulations and the scenarios we studied. As this may be surprising for many readers, we precise in this section where the misunderstandings come from and the underlying phenomenons that explain this apparent paradox.

In this article, we consider *temporary* mitigation policies : the time of intervention is finite and then people return to their normal life. The duration of the intervention may be long, but it is finite. Considering instead infinite time intervention can alter the assessment of a strategy. For instance, suppose that a starting epidemic is annihilated with a drastic mitigation launched at the very beginning when the first people are infected ; then the epidemic never starts again if the mitigations go on forever and if normal life never returns. However, for finite time interventions, the mitigation measures eventually stop. When normal life starts again, the situation is similar to the situation before the mitigation measures, with a naïve population and no immunisation. The epidemic will rise again from the few remaining viruses or from the viruses imported from abroad (see for instance the simulations by Ferguson et al. [8, fig.3] where infections rise after the mitigations are relaxed). The problem has been postponed, rather than solved, by the finite time mitigation.

We consider the mortality in the long run whereas other papers in the literature consider the mortality at a precise date [7]. This may lead to conclusions which are apparently in contradiction, but the results of the two approaches turn out to be compatible once the paradox is understood. Suppose that a first strategy with a rebound of cases after relaxation is settled more early than a second more efficient strategy. The first strategy appears to be preferable if the evaluation occurs before the rebound of cases or when the second strategy has not yet been launched. But if the burden of the epidemic is looked at later, when the efficient late strategy has produced its effects, then the conclusion becomes opposite. This phenomenon explains why Sofonea et al. [7] find good estimates for early mitigations whereas our conclusions are inverse for the long term. As an illustration, we revisit the example of Figure 2. We draw the epidemic with the ratio of removed people *r*(*t*) as a function of *t*. After 26 weeks, the red mitigation seems preferable, but in the long run, the opposite conclusion holds.

If only finite time interventions are allowed, and if assessment is done in the long term, shifting the epidemic with no improvement of the situation is not neutral, it is a waste of resources. If some measures are politically sustainable for 3 months, and if one month is spent in a inefficient set of measures which postpones the problem, then only two months of mitigation policies are left to improve the situation. This explains why in many simulations, early interventions are inefficient in the context of finite time interventions : They are temporal shifts rather than improvements, time is wasted.

### Energy *h* of the system

To make more rigorous the distinction between temporal shift and improvements induced by an intervention, we introduce the energy *h* of the system. A mitigation that lowers *h* ameliorates the situation while a mitigation that lets *h* roughly unchanged acts as a temporal shift. In formula, the energy of a point (*s, r*) is

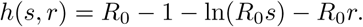

The function *h* is positive for every possible (*s, r*) and minimum at point *P*_*herd*_ = (1*/R*_0_, 1 − 1*/R*_0_) = (*s*_*herd*_, *r*_*herd*_), where its value is 0. We denote by *C*_*h*_ the equienergy curve containing the points (*s, r*) with energy level *h, i*.*e. h*(*s, r*) = *h*. They are the red curves on figure 14. In the absence of mitigation, the energy of the point *M* (*t*) = (*s*(*t*), *r*(*t*)) is unchanged, *i*.*e*. the world *M* (*t*) moves along these red curves. The herd point (*s*_*herd*_, *r*_*herd*_) is drawn in green on the figure. The blue curve on the figure is the trajectory of an epidemic where two mitigations have been launched. During the 2 mitigation periods, the epidemic crosses these level lines, dissipates energy, and the energy *h* in the final situation is smaller than the initial energy. Thus *h* is analogous to energy in physics. It is constant when the system evolves freely (no mitigation), and it diminishes when breaking/mitigation dissipates energy from the system.

**Figure 13:**
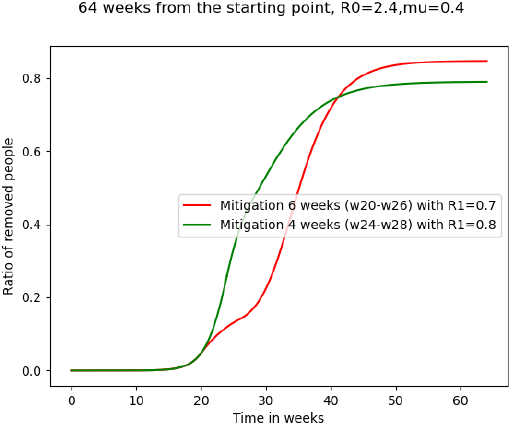
Two mitigations with different plannings.

**Figure 14:**
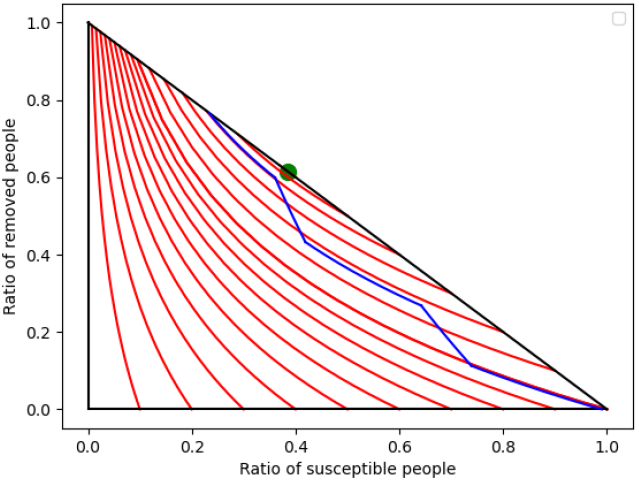
Equienergy curves and a pandemic with 2 mitigations

The geometry of the equienergy curves show that *h* is an indirect measurement of the total burden (past and future, *i*.*e*. including the mortality to come) in the absence of further intervention. Two points on the same equienergy curve go to the same point at infinity if no mitigation measures are set any more : two situations *S*_1_ = *S*(*s*_1_, *i*_1_, *r*_1_) and *S*_2_ = *S*(*s*_2_, *i*_2_, *r*_2_) with the same value of *h* will give the same number *r*_*∞*_ of finally infected people. Moreover, in the absence of further mitigation, the higher the value of *h*, the more people will be infected : *r*_*∞*_ is an increasing function of *h. It follows that the goal of the policy maker is to propose measures that lower h as much as possible before the mitigations are definitively relaxed, with the minimal level of constraint on the population*.

### Variations of *h* during mitigations

How much *h* decreases during a mitigation is governed by the following differential equation. Suppose the epidemic is modelled by a sir system with propagation number *R*_0_ *>* 1 in the absence of mitigation, and by a sir system with propagation number *R*_1_ *< R*_0_ when some restrictions are applied. During the mitigation, *h* is submitted to the differential equation

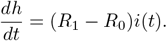

This equation is of particular qualitative importance. Indeed, the goal is to lower *h* rapidly. For a short time *dt*, the variation of *h* is *dh* = (*R*_1_ −*R*_0_)*i*(*t*)*dt*. This means that for a fixed mitigation with number *R*_1_ and a fixed small duration *dt*, the decline of *h* is more important when *i*(*t*) is large. A short time intervention is more efficient when it is applied when the number of infected people *i*(*t*) is large. This result is consistent and may be an explanation for the numerical observation of [8] for an other model :”the majority of the effect of such a [mitigation] strategy can be achieved by targeting interventions […] around the peak of the epidemic.” At the other extreme, if *i*(*t*) = 0, *dh* = 0. We recover the qualitative fact that mitigations applied when *i*(*t*) is very small postpone the problem with no improvement since *h* does not decrease. In particular, an intensive mitigation at the beginning of the epidemic is inefficient.

### On the minimal *r*_*∞*_

Since the ratio of finally infected people *r*_*∞*_ is an increasing function of *h* after the last mitigation, and since *h* is minimal at the herd point, the inequality *r*_*∞*_ *> r*_*herd*_ holds whatever the finite time mitigations. In other words, finite time mitigation measures cannot be used to maintain the epidemic at level zero. In the long term, the minimal share of people that have been infected is at least *r*_*herd*_. Graphically, the limit of the red curves in figure 14 is always located above the herd point *P*_*herd*_.

This impossibility of a zero case strategy by mitigations, or more generally of strategies to reach *r*_*∞*_≤ *r*_*herd*_, is valid only for the finite time strategies considered in this paper. Figure 15 compares a finite time and an infinite time strategy. On the left part, an infinite time strategy is settled and the limit point is below the herd point, *i*.*e. r*_*∞*_ *< r*_*herd*_. On the middle part of the figure, the same mitigation is relaxed after 20 weeks. There is a rebound when relaxing occurs, and *r*_*∞*_ *> r*_*herd*_. On the right part, the mitigation is relaxed after 60 weeks, which makes nearly no difference with 20 weeks, apart from the delay in the 60 weeks case. The sporadic cases that remain active yield quite the same rebound of the epidemic in both cases.

**Figure 15:**
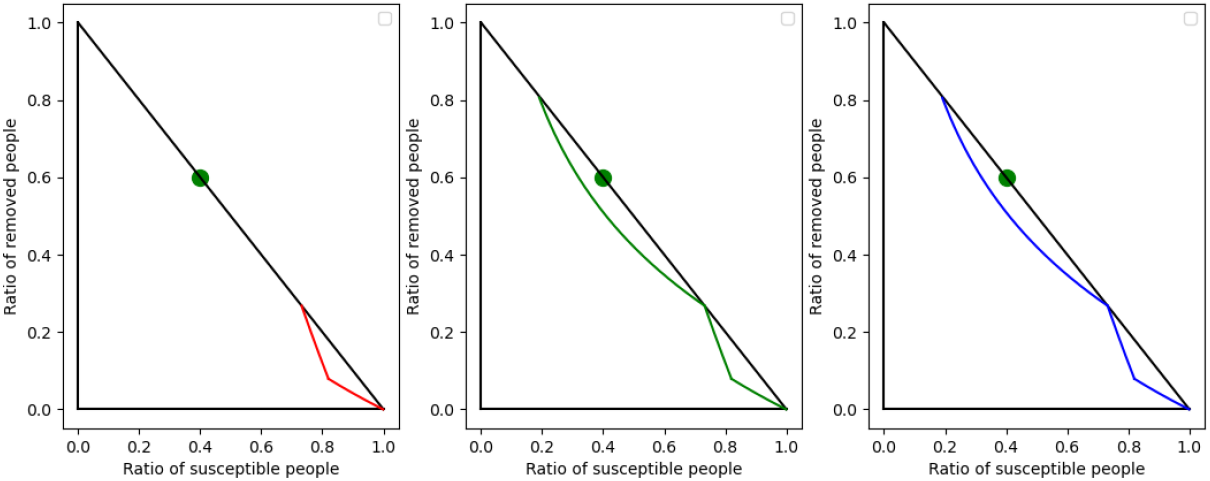
Finite and infinite mitigations

This theoretical result is consistent with on the ground situations for COVID-19. Several countries first tried to develop a zero case strategy, and most of them finally desisted from this strategy [9]. Our analysis suggests a little more : For a given epidemic, it will be difficult if not impossible to maintain *r < r*_*herd*_ in the long run.

The inequality *r*_*∞*_ *> r*_*herd*_ for any finite time strategy is an epidemiological analogue of the mechanical situation with a bike on a sloppy road. If an infinite time breaking is possible, the bike can be stopped in the middle of the slope. In contrast, if the breaking time is finite, the breaks are eventually released, the bike will move to the low point of the road and beyond. In this sense, the herd line *s* = *s*_*herd*_ is the analogue of the bottom of the slope. No finite time breaking can stop the epidemic before it crosses the herd line.

### Dynamics and Inertia of the system after the herd ratio

The analogy with the bike makes it easier to understand the role of the herd immunity threshold, measured equivalently by one of two herd ratios 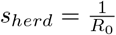 or 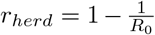. In the vaccine pre-epidemic context, there are no dynamics. The herd ratio *r*_*herd*_ is the proportion of people that need to receive a vaccine to nip in the bud any propagation of the epidemic, preventing its launching from imported cases or remanent cases in the population. In a situation with dynamics, when the epidemic has already started, the situation is different. The epidemic will not stop instantly when the herd ratio is reached. In this dynamical context, *r*_*herd*_ and *s*_*herd*_ still make sense, but have a different interpretation : *i*(*t*) will decrease with time if *s*(*t*) *< s*_*herd*_. In other words, *s*_*herd*_ is the threshold that guarantees the fall of the number of infected people with no mitigation measure. Since *i*(*t*) is proportional to the derivative 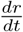, *i*(*t*) is a measure of speed when one tries to minimise the total quantity *r*_*∞*_. The decrease of *i*(*t*) without mitigation after the rational ratio *s*_*herd*_ is the epidemiological counterpart to the fact that a bike that reaches the bottom of the slope will slow down without breaking.

How far the epidemic will go beyond the herd line when mitigations are released is the analogue of the question of how far goes a bike after the bottom of the slope. It depends on the energy *h* of the system. If all the mitigations are released at a point (*s, r*) with energy *h*(*s, r*) = *h*_0_, the share *r* = *r*_*∞*_ of finally infected people at infinity satisfies the equation

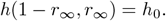

Some comments in the media suggest that once the herd ratio *s* = *s*_*serd*_ is reached, the situation is under control and needs no more supervision. Our models suggest quite the opposite ! Because of the inertia and of the dynamics, it may be necessary to control and slow down the epidemic after the herd ratio is reached. Moreover, in many examples, *i*(*t*) is maximal at the herd ratio *s*(*t*) = *s*_*herd*_. This is true for instance when no mitigation is launched before the herd ratio is reached. In such situations, the equation 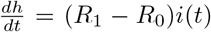 tells that the moment the herd threshold is reached coïncides with maximum effectiveness of the mitigation measures. Thus, not only are the interventions often still necessary when the herd ratio *s* = *s*_*herd*_ is reached, but also, launching the mitigations measures around the herd immunity ratio is good practice according to the sir-model.

### Building mitigations with small *r*_*∞*_

What is the infimum possible *r*_*∞,opt*_ for the number *r*_*∞*_ of finally infected people and what are the strategies to lead to this infimum ? We have seen in the previous section that *r*_*∞*_ *> r*_*herd*_ for any finite time strategy. In this section, we explain that *r*_*∞,opt*_ = *r*_*herd*_, which means that the difference between *r*_*∞*_ and *r*_*herd*_ may be arbitrarily small for a well-designed strategy. We discuss the strategies that lead to this infimum, *i*.*e*. the strategies with *r*_*∞*_ close to *r*_*herd*_.

The naïve and falsely convincing argument that the more restrictive mitigations give the better results on *r*_*∞*_ is wrong. If the mitigation is too intensive, there will be a large rebound in the epidemic. If the mitigation is too loose, the epidemic is not stopped enough. An adequate calibration of the mitigation is thus necessary to get an optimal result, not too intensive to avoid a large rebound, nor too loose to sufficiently dampen the dynamics. This is illustrated in figure 16 with 3 mitigations starting at the same point. The black curve is the epidemic when no mitigation occurs. The mitigation in blue is too loose and the trajectory goes beyond the herd point. The mitigation in green is suitable towards the herd point. The mitigation in red is too strict : a large rebound occurs after the mitigation is stopped. The optimal constant mitigation is understood : its reproduction number is

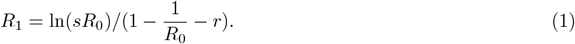

where we suppose that the world is in situation (*s, r*) with *r < r*_*herd*_. Such a mitigation applied an infinite time would lead to the herd point. In practice, the strategy is applied a finite long enough time, the herd point is approached, and *r*_*∞*_ is close to *r*_*herd*_. This is illustrated by the green curve in figure 16 which uses the above formula for *R*_1_.

**Figure 16:**
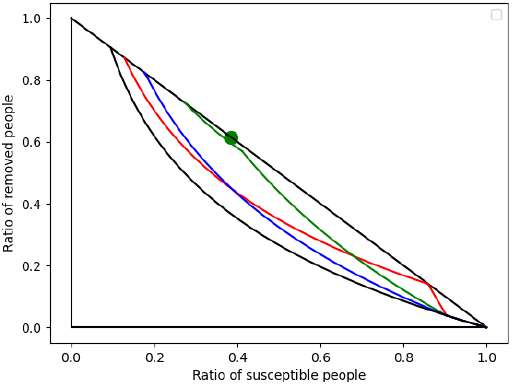
three mitigations : too harsh, correct, and too loose

An other question is the timing. Is it necessary to launch a strategy long before the herd immunity threshold to settle a strategy with *r*_*∞*_ ≃*r*_*herd*_ ? The answer is no from a theoretical point of view. As long as *s*(*t*) *> s*_*herd*_, it is not too to late to launch a strategy that leads to *r*_*∞*_ close to *r*_*herd*_. For instance a mitigation strategy with *R*_1_ as above works. If *s*(*t*) *< s*_*herd*_, the formula implies *R*_1_ *<* 0 which is an impossible physically. In the formula, *R*_1_ tends to zero when *s* tends to *s*_*herd*_, which is a rephrasing of the high intensity required for a late mitigation. This means that on the ground, there are limitations besides theoretical limitations since *R*_1_ cannot be arbitrarily low.

The two questions, intensity and timing, are correlated. There is a large range of possibilities for the start date of an optimal mitigation. The formula for *R*_1_ shows that the later the start date, the more vigorous the intervention must be. But on the other hand, the later the intervention, the shorter the duration of the mitigation for the same result. We can use this correlation between timing and intensity to revisit and explain the initial example of figure 2 or the red mitigation in figure 16. The early mitigation of figure 2 was inefficient because the intensity was not consistent with the starting point. The mitigation was too intensive for its earliness, leading to poor results.

To get order of magnitudes, we illustrate in table 17 possible consistent values for the intensity and duration of the mitigations. We fix *R*_0_ = 3 and *μ* = 0.33. An epidemic rises, and a mitigation is launched when *s* = *s*_*start*_ with the above optimal formula for *R*_1_. The mitigation is relaxed so that *r*_*∞*_ = *l *r*_*herd*_, with *l* = 1.1 or *l* = 1.2. Thus by construction, all scenarios have a burden dependent on *l* but not on the choice of *s*_*start*_ for a fixed *l*. The table reports the values of *R*_1_ and the duration of the mitigation for different values of 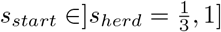.

Remark that there is some flexibility in the choice of *R*_1_ for a fixed *r*_*∞*_. When *R*_1_ is chosen small in the set of possible values, there will be a rebound in the epidemic whereas a large *R*_1_ will yield a strategy with no rebound after the mitigation is relaxed. However, both strategies have the same *r*_*∞*_. Thus the existence of a rebound is not an indicator of an overshoot and of a badly calibrated mitigation.

### Indeterminacy of *R*_0_ and timing implications

Many computations above rely on the estimate of the reproduction number *R*_0_. For instance, the computation of *r*_*herd*_, the energy *h*, the reproduction number *R*_1_ of the optimal mitigation depend on *R*_0_. However the value of *R*_0_ is not well known. Determining *R*_0_ is an important and difficult question. See [10], for the methodology implemented in *Our world in data*, and the many references therein. In this section, we discuss the implications of this indeterminacy on the choice of a strategy. In particular, we exhibit a strategy implementable on the ground without knowing *R*_0_, thus bypassing the difficulty to estimate *R*_0_.

There are many reasons that make it difficult to estimate 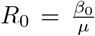. For a virus, *β*_0_, hence *R*_0_, depend on the variants that propagate, they evolve with time. The number *R*_0_ to be used in the modelling can be different from the *R*_0_ that has been computed at the beginning of the epidemic because of the change in the variants. This problem is amplified by the interaction between vaccines and variants. If a vaccine has efficacy *e* with 0 *< e <* 1, and the share of vaccinated people is *r*, then the fraction of the population protected by the vaccine is *re*. These people are to be placed in the removed compartment of the sir system. Thus some continuous exchanges between the *s*-compartment and the *r*-compartment occur along with the evolution of the variants and their interactions with vaccinated people, making the calibration of *R*_0_ difficult.

An other difficulty is that the sir model is valid only locally because of heterogeneity. For a large territory, one can choose a constant *R*_0_ suitable to aggregate the constants of each local area where homogeneity makes sense. However, the local areas evolve independently and the choice of *R*_0_ may lead to a coherent modelling only for a short time. An other heterogeneity was pointed out in [1]. The more the individuals have contacts, the more they spread the virus and the sooner they are infected. Heavy spreaders are in average removed sooner, and *R*_0_ decreases with time. Using the basic reproduction number *R*_0_ computed at the beginning of the epidemic is thus expected to yield overestimation of *r*_*∞*_.

We may formalise these remarks as follows. There is a propagation number *R*_0_(*t*) depending on time, on the (unknown) distribution of heterogeneity, on the immunity evolving with vaccination. For a short enough period of time, all parameters can be seen as constants. The time-varying *R*_0_(*t*) can thus be approximated by a constant and the sir model makes sense.

There exists an optimal intervention which is as late and as strict as possible. If 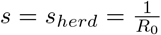 in formula 1, we get *R*_1_ = 0 *independently of the value of R*_0_. This corresponds to a harsh mitigation. Moreover, in the absence of mitigation, *s*(*t*) = *s*_*herd*_ is reached at the moment the number of infected people starts to decrease. Thus there exists an optimal mitigation that starts when the ratio *i*(*t*) of people infected naturally decreases. In contrast to the estimation of *R*_0_ which is very difficult, the moment when *i*(*t*) starts to decrease is easier to monitor. It can be estimated with a detection of the virus in the sewage system. This has already done in France through the “Réseau Obepine” [5].

To sum up, the start time and the intensity of the mitigation should be consistent, and this consistency is difficult to achieve because of the indeterminacy of *R*_0_(*t*) which is a constant only locally in time. This suggests that the following policy independent of *R*_0_ could be considered. The public authorities organise the monitoring of the virus in the sewage system. When *i*(*t*) decreases, it may be interpreted as “the herd immunity ratio is reached”. Then it is a good moment to launch the communication to the public to slow down the spreading and the dynamics with a vigorous mitigation. We think that the feasibility of this proposal needs to be considered as it has several advantages. The mitigation time is shorter than for other intervention times, making the acceptance of the strategy easier. The public confidence in the political decisions is preserved because there are fewer risks of contradictory decisions induced by bad estimates of *R*_0_.

## Data Availability

All data produced in the present work are contained in the manuscript

## 4 Supplementary

### 4.1 Description of the model

In this section, we describe the sir-controlled model that we use throughout the article, which involves some non continuity considerations that need to be clearly formulated.

We consider an epidemic starting at time *t* = 0. Three functions *s, i, r* of the time *t* are considered : *s*(*t*) is the share of people not infected up to *t, i*(*t*) is the share of people infected at *t* and *r*(*t*) is the share of people who were infected before *t*, but are cured at *t*. By construction, for every non negative *t, s*(*t*) + *i*(*t*) + *r*(*t*) = 1. We do not consider births in the model. The classical sir equations are:

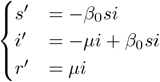

where

- *μ* is a strictly positive constant and depends on the epidemiological context. It governs the speed at which infected people are removed.
- *β*_0_ is a strictly positive constant and 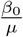 is the initial propagation number, *i*.*e*. the average number of infections originated from the first infected persons. We use the classical notation 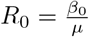.

#### Remark 1.

*The classical SIR-system is often presented using absolute numbers, whereas the above version considers ratios rather than sizes of populations. For instance, in [11], the Kermack-McKendrick SIR epidemic model is presented with the number I of infected people, and N the size of population, whereas we use the share* 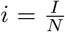 *instead. We use the lowercase notation* **sir-system** *rather than the uppercase notation SIR-system as a reminder of this choice*.

When mitigation policies apply, *β*_0_ is not a constant any more. We consider a model where we replace the constant *β*_0_ with a function *β* = *β*(*t*) depending on the time *t*. When no mitigation strategy is set up, *β*(*t*) = *β*_0_. When mitigation strategies occur, 0≤ *β*(*t*) *< β*_0_. The condition *β*(*t*) *> β*_0_ is mathematically possible, but corresponds to people gathering and transmitting the virus more than expected, thus is hardly realistic.

On the other hand, *μ* is constant as before and is independent of the mitigation strategy.

Summing up, our model to include mitigation strategies is a derivation of the classical sir model where *β*_0_ is replaced by a non negative function *β* = *β*(*t*). In mathematical terms :

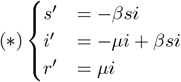

In the following, we will use the expression “sir-controlled model” for this model, or sometimes only “sir-model” for simplicity, assuming it is implicit and clear that *β* is not a constant.

When a mitigation is launched at time *t*, a discontinuity occurs for the function *β*. Thus we need to consider non continuous functions *β*(*t*) and we suppose only that *β*(*t*) is piecewise continuous on [0, + *∞*[. In this non continuous context, we define a solution of a controlled sir-system as follows.

#### Definition 2.

*We suppose that there exists a subdivision a*_0_ = 0 *< a*_1_ *< … < a*_*k*−1_ *< a*_*k*_ = +*∞ such that β is continuous on each*]*a*_*j*_, *a*_*j*+1_[. *Moreover, we suppose that for j* ∈ {0, …, *k* − 1}, *the right limit* 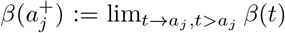 *and for j* ∈ {0, …, *k* − 2}, *the left limit* 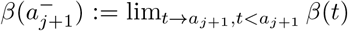 *exist. In general*, 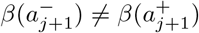.

*Let* 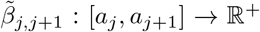 *be the continuous function that extends β* :]*a*_*j*_, *a*_*j*+1_[→ *ℝ*^+^. *A solution of the sir controlled system is a triple of functions* (*s, i, r*) *defined on* [0, +*∞*[*satisfying*

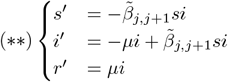

*on each bounded interval* [*a*_*j*_, *a*_*j*+1_] *for j* ≤ *k* − 2 *and on the last unbounded interval* [*a*_*k*−1_, +*∞*[. *In particular, s has a left and right derivative at each point a*_*i*_, *i* ∈ {1, …, *k* − 1} *and a derivative at a*_0_.

*We define* 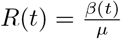. *If β*(*t*) = *β*_0_ *for all t, then R*(*t*) = *R*_0_ *is the classical propagation number*.

### 4.2 Existence of solutions

In this section, we prove that under our hypothesis for *β*, the sir controlled systems have a unique solution, with regularity and monotony properties. These are standard facts when *β* is a constant(see for instance [11]) and the proof extends easily when *β* is a piecewise continuous function.

#### Theorem 3.

*Let* (*s*_0_, *i*_0_, *r*_0_) *with s*_0_ *>* 0, *i*_0_ *>* 0, *r*_0_ ≥ 0 *and s*_0_ + *i*_0_ + *r*_0_ = 1. *Then there exists a unique solution* (*s, i, r*) *of the sir-controlled system* (*\*\**) *defined on* [0, +*∞* [*satisfying* (*s*(0), *i*(0), *r*(0)) = (*s*_0_, *i*_0_, *r*_0_). *Moreover, the solution satisfies the following properties:*

- *s, i, r are continuous and their restrictions on the intervals* [*a*_*j*_, *a*_*j*+1_], *j* ∈{0, …, *k*− 2} *and* [*a*_*k*−1_, + *∞* [*are C*^1^,
- ∀*t* ≥ 0, *s*(*t*) *>* 0 *and i*(*t*) *>* 0,
- ∀*t >* 0, *r*(*t*) *>* 0,
- *r is strictly increasing and s is decreasing*,
- ∀*t* ≥ 0, *s*(*t*) + *i*(*t*) + *r*(*t*) = 1,
- *The limits s*_*∞*_ := lim_*t*→*∞*_ *s*(*t*), *i*_*∞*_ := lim_*t*→*∞*_ *i*(*t*) *and r*_*∞*_ = lim_*t*→*∞*_ *r*(*t*) *exist*.
- *i*_*∞*_ = 0, *s*_*∞*_ + *r*_*∞*_ = 1.

*Proof*. We first consider the equations on the interval [*a*_0_ = 0, *a*_1_]. Let *I* ⊂ [*a*_0_, *a*_1_] be the maximal interval where the solution with initial condition (*s*(0), *i*(0), *r*(0)) = (*s*_0_, *i*_0_, *r*_0_) is defined.

If *s*(*t*) = 0 for some *t* ∈*I*, then *s*(*t*) = 0 for all *t* ∈*I* since *s* satisfies a first order linear equation *s′* = (−*βi*)*s*, contradicting the value of *s*(0). Similarly, *i*(*t*) cannot vanish. Thus *i*(*t*) *>* 0 and *s*(*t*) *>* 0. It follows by derivation that *r* is strictly increasing and *r*(*t*) *>* 0 for *t >* 0, and that *s* is decreasing. By the sir-system, *s* + *i* + *r* is a constant function since its derivative is zero, and its initial value is 1.

Let *b* = *sup*(*I*), *i*.*e. I* = [*a*_0_, *b*[or *I* = [*a*_0_, *b*] with *b*≤ *a*_1_. The limits *s*(*b*^−^), *i*(*b*^−^), *r*(*b*^−^) at *b* exist since *s* is positive decreasing, *r* is increasing bounded by 1, and *s* + *i* + *r* = 1.

If *b* = +*∞*, it remains to prove that *i*_*∞*_ = 0. If *i*_*∞*_ *>* 0, the equation *r′* = *μi* shows that *r*_*∞*_ = +*∞*, contradiction. Thus *i*_*∞*_ = 0 and *r*_*∞*_ + *s*_*∞*_ = 1 follows.

If *b <* +*∞*, the limits of the derivatives *s′*(*b*^−^), *i′*(*b*^−^), *r′*(*b*^−^) exist by the sir-system and the limits of *s, i, r*.

Therefore, the solution can be defined at *b*. By maximality of *I*, it follows that *b* = *a*_1_. We conclude by induction, replacing *t* = *a*_0_ by *t* = *a*_1_, with fewer intervals for the definition of *β*.

#### Proposition 4.

*Let a sir-system with β*(*t*) = *β*_0_ *for t* ≥ *t*_0_. *If* 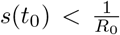, *then i*(*t*) *is strictly decreasing for t* ≥ *t*_0_. *If* 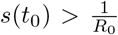, *then there exists a t*_1_ *> t*_0_ *such that i is strictly increasing on* [*t*_0_, *t*_1_] *and strictly decreasing on* [*t*_1_, +*∞*[. *Moreover t*_1_ *is characterised by* 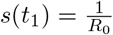.

*Proof*. If 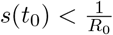, then the function *β*_0_*s* − *μ* is negative at *t*_0_. As *s* is decreasing, the same holds for *t* ≥ *t*_0_. By the sir-system, *i′* = (*β*_0_*s* − *μ*)*i* and then *i′*(*t*) *<* 0 for *t* ≥ *t*_0_.

If 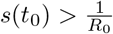, then by the same equation of the sir-system, *i* is at first increasing. Since *i*_*∞*_ = 0, the function *i* must have a maximum at some *t*_1_ *> t*_0_. At this point *i′*(*t*_1_) = *β*_0_*s*(*t*_1_) − *μ* = 0, *i*.*e*. 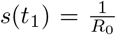. Since *s* is decreasing, *i* is at first increasing, reaches its maximum when 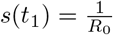 and then is decreasing. The increase and the decrease of *i* are strict, otherwise *i, s*, and *r* = 1 −*i* −*s* are constant on some interval, whereas *r* is strictly increasing.

#### Corollary 5.

*If a mitigation is stopped at time t*_1_ *with s*(*t*_1_) *> s*_*herd*_, *then i will be increasing for t > t*_1_ *during a certain time. In other words, a new wave is rising*.

### 4.3 Foliation of the triangle and solutions of the sir-system

Our approach to study the solutions of the sir-system is to work in the plane ℝ^2^ (rather than ℝ^3^) with coordinates (*s, r*), forgetting the quantity *i* = 1− *s* −*r*. When *β*(*t*) is a constant function, the trajectories of the solutions in ℝ^2^ are included in a set with equation *H*(*s, r*) = *c* for some function *H* and some constant *c*. The coordinates (*s, r*) lies in a triangle *T* and the loci *H*(*s, r*) = *c* when *c* varies draw a foliation on *T*. In this section, we introduce the foliation and we study its geometry (Theorem 7). We then derive the consequences of this geometrical study on the solutions of the sir-system associated to *β* (Proposition 10).

#### Definition 6.

*Let T* ⊂ *ℝ*^2^ *be the set of points* (*s, r*) *with s >* 0, *r* ≥ 0 *and s* + *r* ≤ 1. *Let R* ≥ 0 *and H* :]0, +*∞*[*×ℝ* → *ℝ with H*(*s, r*) = ln(*s*) + *Rr. If R* ≥ 1, *we let* 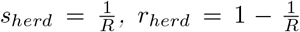, *i*_*herd*_ = 0, *M*_*herd*_ = (*s*_*herd*_, *r*_*herd*_). *A R-leaf C*_*c*_ *is a set in T defined by C*_*c*_ := {*M* ∈ *T, s*.*t. H*(*M*) = *c*} *for some constant c. We let i be the function on T defined by i*(*M*) = 1 − *s*(*M*) − *r*(*M*) *where s, r are the coordinate functions*.

The leaves for *R* = 2.6 and *R* = 0.8 are shown in figure 18. Later on, we will use this foliation for *R* = *R*_0_ the basic reproduction number but also for *R* = *R*_1_, the reproduction number induced by some mitigation.

**Figure 17:**
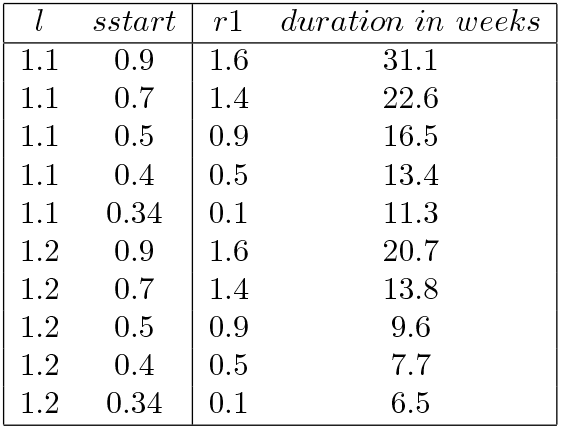
R1 and duration of the mitigation

**Figure 18:**
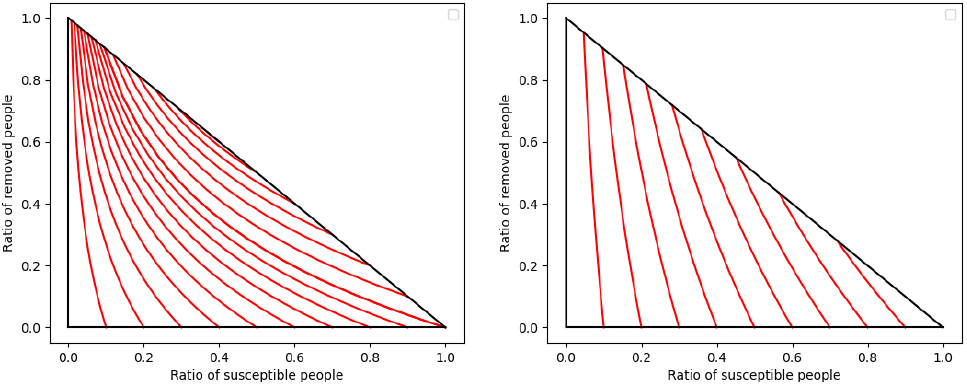
Leaves for *R* = 2.6 and *R* = 0.8

#### Theorem 7.

*1. The R-leaves C*_*c*_ *form a foliation of T, which means for us that the leaves C*_*c*_ *are smooth and connected. Moreover, the leaves are compact*.

*2. If R* ≥ 1, *the point M*_*herd*_ *is a leaf reduced to a point. The other leaves are curves*.

*3. If R* ≤ 1, *M* = (1, 0) *is a leaf reduced to a point. The other leaves are curves*.

*4. For any leaf C*_*c*_, *there exists a unique point M*_*∞*_ *on C*_*c*_ *such that r*(*M*_*∞*_) = max{*r*(*M*), *M* ∈ *C*_*c*_}. *Moreover, s*(*M*_*∞*_) = min{*s*(*M*), *M* ∈ *C*_*c*_} *and i*(*M*_*∞*_) = 0 *(When precision is required, we denote M*_*∞*_ = *M*_*∞*_(*C*_*c*_)).

*5. For any leaf C*_*c*_ *with c* ≥ 0,*there exists a unique point M*_*init*_ *on C*_*c*_ *(sometimes denoted M*_*init*_(*C*_*c*_) *) such that r*(*M*_*init*_) = min{*r*(*M*), *M* ∈ *C*_*c*_}. *Moreover, s*(*M*_*init*_) = max{*s*(*M*), *M* ∈ *C*_*c*_} *and i*(*M*_*init*_) = 0.

*6. If a leaf C*_*c*_ *is non empty for c >* 0, *then R >* 1.

*7. A point M* ∈ *T is a point M*_*∞*_ *of some leaf C*_*c*_ *if and only if i*(*M*) = 0 *and* 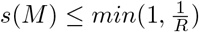.

*8. Let C*_*c*_ *be a leaf. If C*_*c*_ *is reduced to a point, then T \ C*_*c*_ *is connected, otherwise T \ C*_*c*_ *has 2 components*.

*Proof*. The function *H*(*s, r*) increases strictly when *s* increases. Hence in order to study the maximum of *H*, we can restrain it to the diagonal {(*s, r*), *s >* 0, *r* ≥ 0, *s* + *r* = 1}, or equivalently we study the function *H*(*s*, 1 − *s*) on the interval]0, 1]. The derivative 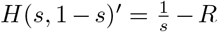. shows that the maximum of *H* is obtained for 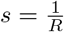 if *R* ≥ 1 and for *s* = 1 is *R <* 1. It follows that the maximum of *H* is realised on a unique point of *T* which is a leaf reduced to a point *M*_*max*_. If *R* ≥ 1, *M*_*max*_ = *M*_*herd*_, otherwise, *M*_*max*_ = (1, 0).

Let *M*_1_ = (*s*_1_, *r*_1_) and *M*_2_ = (*s*_2_, *r*_2_) be two points on the same leaf *C*_*c*_ with *r*_2_ *< r*_1_. The graph Γ of the function[*r*_2_, *r*_1_] → *ℝ, r* → *s* = *e*^*c*−*Rr*^ is included in the locus *H* = *c* and contains *M*_1_ and *M*_2_. To prove the connectedness of *C*_*c*_, it remains to prove that Γ ⊂ *T*. By convexity of the exponential function, Γ lies below the segment [*M*_1_, *M*_2_] and by monotony, Γ is included in the rectangle [*s*_1_, *s*_2_] *×* [*r*_2_, *r*_1_]. The part of the rectangle below [*M*_1_, *M*_2_] is the triangle *Conv*(*M*_1_, *M*_2_, *M*_3_) with *M*_3_ = (*s*_1_, *r*_2_). By convexity of *T*, it follows that Γ ⊂ *Conv*(*M*_1_, *M*_2_, *M*_3_) ⊂ *T*.

A point (*s, r*) ∈ *C*_*c*_ satisfies *r* ≤ 1 hence *s* ≥ := *e*^*c*−*R*^. Thus *C*_*c*_ is compact as the intersection of the closed set ln(*s*) + *Rr* = *c* with the compact *T* ∩ {*s* ≥ }. It follows by compacity and connectedness that *r*(*C*_*c*_) = [*r*_1_, *r*_2_] for some *r*_1_, *r*_2_ ∈ [0, 1] and that *C*_*c*_ is the graph Γ of *r* ∈ [*r*_1_, *r*_2_] → *s* = *e*^*c*−*rR*^. The smoothness of *C*_*c*_ follows.

The existence and uniqueness of *M*_*∞*_ and *M*_*init*_ in items 4 and 5 follow from the description of *C*_*c*_ as a graph of a monotonous function of *r* in the previous paragraph. The condition *i*(*M*_*∞*_) = 0 holds : otherwise, *M*_*∞*_ is in the interior of the triangle *T* and there would exist *r*_3_ *> r*_2_ = *r*(*M*_*∞*_) with 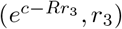 in *T*, contradicting the maximality of *r*(*M*_*∞*_) on the leaf. Similarly, *M*_*init*_ exists by compacity and satisfies the maximality conditions by monotony. If *c* = 0, then obviously *M*_*init*_ = (1, 0) and point 5) is true. If *c >* 0, we have *r*(*M*_*init*_) *>* 0. If *i*(*M*_*init*_) ≠ 0, then *M*_*init*_ is in the interior of the triangle *T* and we contradict the minimality of *r* as above.

If *R* ≥ 1, ln(*s*) + *Rr* ≤ ln(*s*) + *R*(1 − *s*) ≤ 0 for *s* ≤ 1. This proves the sixth item.

By construction, a point *M*_*∞*_ is a point on a *R*-leaf with *r*(*M*) maximum and we know that *i*(*M*_*∞*_) = 0. Thus a point *M* ∈ *T* is a point *M*_*∞*_(*C*_*c*_) for some *c* if and only if *i*(*M*) = 0 and *r*(*M*) = max{*r*(*M′*)} where *M′* runs through the points in *T* satisfying *i*(*M′*) = 0 and *H*(*M′*) = *c*. Rephrasing informally, the points *M*_*∞*_ are the leftmost points of *T* on the segment *i* = 0 among those *M′* which have the same *H*-value.

For item 7), if *R <* 1, we need to show that any point *M* with *i*(*M*) = 0 is equal to *M*_*∞*_(*C*_*c*_) for some *c*. We let *c* = *H*(*M*). The function *s* → *H*(*s*, 1 − *s*) = ln(*s*) + *R*(1 − *s*) is strictly increasing, thus *M* is the unique point *M* with *i*(*M*) = 0 and *H*(*M*) = *c*. Therefore *M* = *M*_*∞*_(*C*_*c*_).

If *R* ≥ 1, the function *s* → *H*(*s*, 1 − *s*) = ln(*s*) + *R*(1 − *s*) is strictly increasing from *s* = 0 to 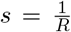, then strictly decreasing from 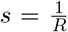 to *s* = 1. If *c* := *H*(*M*) *< H*(1, 0) = 0 or if 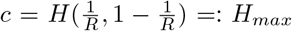, then as above *M* is the unique point with *i*(*M*) = 0 and *H*(*M*) = *c*. Therefore *M* = *M*_*∞*_(*C*_*c*_). If *c* ≥ 0 and *c < H*_*max*_, then there are two values *s*_0_, *s*_1_ solutions of *H*(*s*, 1 − *s*) = *c*. The smallest value *s*_0_ satisfies 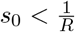 while 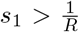. Thus *M*_*∞*_ (*C*_c_) = (*s*_*0*_, 1 − *s*_0_) and *M*_*init*_ (*C*_c_) = (*s*_1_, 1 − *s*_*1*_). It follows that the points *M*_*∞*_ are the points 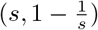 with 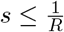.

We have exhibited some leaves that are reduced to points. We show now that the other leaves are curves. We know that a leaf *C*_*c*_ is a graph of a function *r* ∈ [*r*_1_, *r*_2_] → *e*^*c*−*Rr*^ = *s* parameterised by the coordinate *r*. In particular, *C*_*c*_ is reduced to a point if *r*_1_ = *r*_2_ and *C*_*c*_ is a curve otherwise. In particular, if *C*_*c*_ contains at least two different points, it is a curve.

- If *R <* 1, then *c* ≤ 0, and *C*_*c*_ contains the points *M*_*∞*_(*C*_*c*_) and (*e*^*c*^, 0). If *c <* 0, the 2 points are different and *C*_*c*_ is a curve. If *c* = 0, we have already seen that *C*_0_ is a point.
- If *R* ≥ 1 and *c <* 0, we proceed as above. If *R* ≥ 1, and *c* ∈ [0, *H*_*max*_[, then *C*_*c*_ contains the two distinct points *M*_*∞*_(*C*_*c*_) and *M*_*init*_(*C*_*c*_) and it is a curve.
- In the last case *R* ≥ 1 and *c* = *H*_*max*_, we have already proved that *C*_*c*_ = {*M*_*herd*_} is a point.

For the last item, each leaf *C*_*c*_ is connected, thus the connectedness problem reduces to connect the various leaves *C*_*c*_, or equivalently the various points *M*_*∞*_(*C*_*c*_). The set of points *M*_*∞*_(*C*_*c*_) is the set {(*s*, 1 −*s*), *with* 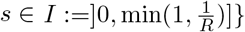. Thus it is connected since *I* is connected.

When a curve *C*_*c*_ is removed from *T, I* is replaced with *J* = *I \* {*s*(*M*_*∞*_(*C*_*c*_))}. If *C*_*c*_ is a point, then by the above, 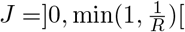 is connected. Otherwise, 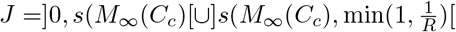, and there are two components *H > c* and *H < c*.

#### Remark 8.

*The standard definition of a foliation is slightly different from the one used in the theorem*.

#### Definition 9.

*Consider a controlled sir system defined for t in an interval I. Let* (*s, i, r*) : *I* → *ℝ*^3^ *be a solution. Let M* (*t*) = (*s*(*t*), *r*(*t*)). *The set τ* ⊂ *ℝ*^2^ *defined by τ* := {*M* (*t*), *t* ∈ *I*} *is called a trajectory*.

*If I* = [*t*_0_, *t*_1_], *t*_1_ *> t*_0_, *or I* = [*t*_0_, +*∞*[, *M*_0_ ∈ *ℝ*^2^ *and* (*s*(*t*_0_), *i*(*t*_0_), *r*(*t*_0_)) = (*s*(*M*_0_), *i*(*M*_0_), *r*(*M*_0_)), *then τ is called a trajectory with initial condition M*_0_.

*The trajectory is constant if τ is reduced to a point M*_*τ*_. *A constant trajectory is stable (or equivalently, the point M*_*τ*_ *is stable) if for every >* 0, *there exists δ such that for every M*_0_ *with* ||*M*_0_ − *M*_*τ*_ || *< δ, the solution M* (*t*) *with initial condition M*_0_ *at t* = *t*_0_ *satisfies* ||*M* (*t*) − *M*_*τ*_ || *< for all t* ≥ *t*_0_.

The following proposition characterises the trajectories in terms of the leaves of the foliation. Essentially, the leaves are unions of trajectories.

#### Proposition 10.

*Suppose that the function β in the sir-controlled system is constant. Let* 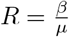 *and consider the foliation associated to R*.

1. *Any trajectory is included in a R-leaf C*_*c*_.
2. *A subset τ* ⊂ ℝ^2^ *is a non constant trajectory if and only if the following conditions are satisfied:*
  - *There exists a R-leaf C*_*c*_ *with τ* ⊂ *C*_*c*_,
  - 0 ∉ *i*(*τ*),
  - *The set r*(*τ*) *is an interval I*_*r*_ *non reduced to a point*.
3. *A trajectory with initial condition M*_0_ *is constant if and only if i*(*M*_0_) = 0. *A constant trajectory is stable if and only if R <* 1 *or (R* ≥ 1 *and* 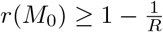.
4. *Let τ* = {*M* (*t*)} *be a non constant trajectory. Recall that* (*s*_∞_, *r*_∞_) = lim_*t*→∞_ *M* (*t*) *is well defined. Let c* = ln(*s*_∞_) + *Rr*_∞_ *and let C*_*c*_ *be the associated leaf. Then M*_∞_(*C*_*c*_) = (*s*_∞_, *r*_∞_). *In other words, the two notions of point at infinity (in the foliation and in the differential equation) coïncide*.

*Proof*. It follows from the sir-system that the equation 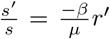. By integration, it follows that *H*(*s, r*) = ln(*s*) + *Rr* is constant on a trajectory. This proves the first item.

For item 4), we have seen that *s*_∞_ + *r*_∞_ = 1 and that *C*_*c*_ is closed. It follows that (*s*_∞_, *r*_∞_) ∈ (*C*_*c*_ ∩ (*i* = 0)). Now, *C*_*c*_ has one ore two points *M* with *i*(*M*) = 0, namely *M*_∞_(*C*_*c*_) and maybe *M*_*init*_(*C*_*c*_). However, *r*(*t*) is strictly increasing, thus *M*_*init*_ defined by the minimality of *r* cannot be equal to (*s*_∞_, *r*_∞_). By elimination, (*s*_∞_, *r*_∞_) = (*M*_∞_(*C*_*c*_)).

For the point 2), it is easy to show that a trajectory *τ* satisfies these three conditions. Conversely, suppose that *τ* satisfies the three conditions. We consider first the case with *I*_*r*_ = [*a, b*] is a closed interval. Remark that any point *M* of *τ* is determined by the value *r*(*M*), namely *M* = (*e*^*c*−*Rr*(*M*)^, *r*(*M*)). Let *M* (*t*) be the solution defined on *I* = [0, +∞[and with initial condition *M* (0) = (*e*^*c*−*Ra*^, *a*). Then *b* ≤ *r*(*M*_∞_(*C*_*c*_)) = *r*_∞_ by construction of *M*_∞_(*C*_*c*_) which has the maximum possible *r* on a *R*-leaf. Moreover, *b < r*_∞_, otherwise *τ* contains *M*_∞_ which is a point with *i* = 0. Thus, by intermediate value theorem, there exists *t*_0_ ∈ [0, +∞[with *r*(*M* (*t*_0_)) = *b*. The set {*M* (*t*), *t* ∈ [0, *t*_0_]} is *τ*. Thus we have proved item 2) when *I*_*r*_ = [*a, b*]. If *I*_*r*_ =]*a, b*[is open, or semi-open, we glue the solutions defined on [*a*_*n*_, *b*_*n*_] with *a < a*_*n*_ *< b*_*n*_ *< b*.

For item 3), it is obvious that the trajectory is constant iff *i*(*M*_0_) = 0. We prove the stability statements in the case *R >* 1 (the case *R* ≤ 1 is easier). When *R >* 1, there are two type of points in *T* satisfying *i* = 0, namely the points *M* of the form *M*_∞_ which are characterised by 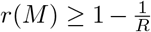, and the points *M* of the form *M*_*init*_ which are characterised by 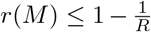. Thus, to prove item 3) when *R >* 1, we need to show that the points *M*_∞_ are stable and that the remaining points are unstable. (If *R* ≤ 1, all points with *i* = 0 are points of the form *M*_∞_).

Let *M*_0_ be any point in *T, M*_0_(*t*) the solution of the sir-system with initial condition *M*_0_, and *M*_0_(∞) = (*s*_∞_, *r*_∞_) be the limit at infinity of *M*_0_(*t*).

Let *D* ⊂ *T* be the locus defined by *i* = 0, let *D*^+^ ⊂ *D* be the set of points (*s, r*), with 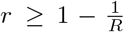, *D*^−^ ⊂ *D* defined by 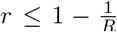, *T* ^*′*^ ⊂ *T* defined by *i* ≠ 0. Thus *T* = *T* ^*′*^ ∪ *D*^+^ ∪ *D*^−^ and *D*^+^ ∩ *D*^−^ is the point 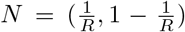. In particular, a function *L* : *T* → *T* is continuous on *T* ^*′*^ ∪ *D*^+^ if its restrictions *L*_1_ : *T* ^*′*^ ∪ *D*^+^ → *T* and *L*_2_ : *D*^−^ → *T* are both continuous (Reason: *L* is continuous on the interior of *T* ^*′*^ ∪ *D*^+^ and on *N* which is in both sets of the covering *T* = (*T* ^*′*^ ∪ *D*^+^) ∪ *D*^−^). We want to apply this remark when *L* : *T* → *T, M*_0_ → *M*_0_(∞) is the limit function. The restriction of *L* to *D*^−^ is identity. Thus we need only prove that the restriction to *T* ^*′*^ ∪ *D*^+^ is continuous. The function *M*_0_ → *H*(*M*_0_(∞)) is continuous because *H*(*M*_0_(∞)) = *H*(*M*_0_). Now the restriction 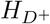 of *H* to the interval *D* is continuous and injective, thus an homeomorphism on its image, with inverse *H*^−1^. By composition, the limit function *L* : *T* ^*′*^ ∪ *D*^+^ → *D*^+^, 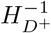 is continuous on *T* ^*′*^ ∪ *D*^+^.

By continuity of *L* on *T* ^*′*^ ∪ *D*^+^, if *M*_0_ ∈ *T* is close to a fixed *M*_∞_ ∈ *D*^+^, *L*(*M*_0_) = *M*_0_(∞) is close to *L*(*M*_∞_) = *M*_∞_. Since for every *t, M*_0_(*t*) is included in the rectangle with diagonal [*M*_0_, *M*_0_(∞)], it follows that *M*_0_(*t*) is close to *M*_∞_ too. This proves the stability of *M*_∞_.

A point *M*_*init*_ in *D\ D*^+^ satisfies 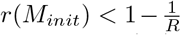. It is unstable since for any choice of the initial condition *M*_0_ close to *M*_*init*_ with *i*(*M*_0_) *>* 0, the limit *M*_0_(∞) satisfies *r*(*M*_0_(∞)) ≥ 1 − 1*/R*. Thus for *t* large, *M*_0_(*t*) is not close to *M*_*init*_.

### 4.4 Optimal mitigations

The space of all possible mitigations is infinite dimensional. Among all possible mitigations, what is the optimal value *s*_*opt*_ maximising the number *s*_∞_ of never infected people ? The goal of this section is to compute this optimal value (Theorem 12). This result holds for finite time controls, *i*.*e*. life goes back to normal after some time and mitigations may be arbitrarily long but don’t last forever. The proof is a direct consequence of our study of the stability of fixed points.

#### Definition 11.

*A controlled sir system has a finite time control β if β*(*t*) = *β*_0_ *for t large enough*.

*We fix an initial point M*_0_ = (*s*_0_, *r*_0_) *with i*(*M*_0_) ≠ 0, *and we denote by s*_∞_(*β*) *the limit s*_∞_ *of the sir system. We define s*_*opt*_ = sup_*β*_ *s*_∞_(*β*), *where β runs through all finite time controls*.

#### Theorem 12.

*With the notations of definition 11*,

- *If s*_0_ ≥ *s*_*herd*_, *then s*_*opt*_ = *s*_*herd*_.
- *If s*_0_ ≤ *s*_*herd*_, *then s*_*opt*_ = *s*_0_.

*Proof*. First, consider the case *s*_0_ ≤ *s*_*herd*_. Since *s*(*t*) is decreasing and *s*(0) = *s*_0_, we have for any *β, s*_∞_(*β*) ≤ *s*_0_ and then *s*_*opt*_ ≤ *s*_0_. If we take *β* = 0, then lim_*t*→∞_ *M* (*t*) = (*s*_0_, 1 − *s*_0_), which is a stable limit by proposition 10, item 3. Let now *β*^*′*^ defined from *β* by relaxation after some time *t*_*r*_ to get a finite time strategy: *β*^*′*^(*t*) = 0 for *t* ∈ [0, *t*_*r*_] and *β*^*′*^(*t*) = *β*_0_ for *t > t*_*r*_. If *t*_*r*_ is large, then *M* (*t*_*r*_) is as close as we want to (*s*_0_, 1 − *s*_0_). By stability, it follows 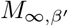 is as close as we want to (*s*_0_, 1 − *s*_0_). This shows that *s*_*opt*_ ≥ *s*_0_.

Suppose now that *s*_0_ ≥ *s*_*herd*_. For any finite time strategy *β*, there exists a time *t*_0_ such that *β*(*t*) = *β*_0_ for *t* ≥ *t*_0_. For *t* ≥ *t*_0_, we can apply proposition 10, item 4 and Theorem 7, item 7, to conclude that *s*_∞_(*β*) ≤ *s*_*herd*_. Thus, *s*_*opt*_ ≤ *s*_*herd*_.

Take *R*_1_ in order to have the relation: ln *s*_0_ + *R*_1_*r*_0_ = ln *s*_*herd*_ + *R*_1_*r*_*herd*_. Explicitly: 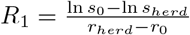. Observe that we are in a realistic situation : *R*_1_ ≥ 0 and *R*_1_ ≤ *R*_0_. This follows from the hypothesis *R*_0_*s*_0_ ≥ 1 and from the inequalities *r*_*herd*_ − *r*_0_ *> s*_0_ − *s*_*herd*_, ln *x < x* − 1 for *x >* 1. Take the solution (*s*(*t*), *i*(*t*), *r*(*t*)) of the sir system with *β*(*t*) a constant function equal to *β*_1_ = *μR*_1_ and with initial condition (*s*_0_, 1 − *s*_0_ − *r*_0_, *r*_0_). The choice of *R*_1_ implies that *s*_∞_ = *s*_*herd*_ and *r*_∞_ = *r*_*herd*_. Now, we relax at a time *t*_*r*_, *i*.*e*. we consider *β*^*′*^(*t*) = *β*(*t*) for *t* ≤ *t*_*r*_ and *β*^*′*^(*t*) = *β*_0_ for *t < t*_*r*_. Then *M* (*t*_*r*_) is arbitrarily close to *M*_*herd*_ if *t*_*r*_ is large enough, and since *M*_*herd*_ is a stable point, *M*_∞_(*β*^*′*^) remains as close as we want to *M*_*herd*_. This shows that *s*_*opt*_ ≤ *s*_*herd*_.

#### Remark 13.

*The theorem states that there exists a mitigation with s*_∞_ *as close as we want to s*_*opt*_, *but there is no finite time strategy with s*_∞_(*β*) = *s*_*opt*_.

### 4.5 Energy of the system

In this section, we build an analogy with the mechanical systems. We introduce the energy function *h*_0_ which is a renormalisation of *H* when *R* = *R*_0_. We show that the number of finally infected people *r*_∞_ is an increasing function of *h*_0_ (Theorem 16). This is analogous to the fact that a collision between highly energetic objects implies serious damage. The breaking mechanical power (in the usual physical understanding) of the mitigation is then 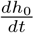. The formula of Theorem 17 shows that this power is negative for a mitigation. Thus a mitigation can be seen as a breaking which lowers the energy and the final damage. When *i* = 0, the power is zero. Thus breaking when *i* is small is not efficient.

#### Definition 14.

*Recall the function H*(*s, r*) = ln(*s*)+*Rr. When the constant R is equal to the basic propagation number R*_0_ *of an epidemic, we use the specific notation H*_0_(*s, r*) = ln(*s*) + *R*_0_*r for the function H. We let h*_0_(*s, r*) = −*H*_0_(*s, r*) − ln(*R*_0_) + *R*_0_ − 1. *The quantity h*_0_(*s, r*) *is called the energy at point* (*s, r*).

#### Proposition 15.

*If R*_0_ ≥ 1, *the energy h*_0_ *is non negative on the triangle T and* min_(*r,s*)∈*T*_ *h*_0_(*r, s*) = 0. *Moreover M*_*herd*_ *is the unique point with h*_0_(*M*_*herd*_) = 0. *If R*_0_ *<* 1, *then* (1, 0) *is the unique point of T where the energy is minimal*.

*Proof*. We have proved in Theorem 7, first paragraph, that *H*_0_ has a unique maximum on *T* located at *M*_*herd*_ or (1, 0) depending on the value of *R*_0_. The result for *h*_0_ follows.

#### Theorem 16.

*Let M*_0_ = (*s*_0_, *r*_0_) ∈ *T with i*(*M*_0_) *≠* 0. *Let M* (*t*) *be the solution of the non controlled sir-system with β*(*t*) = *β*_0_ *and initial condition M*_0_. *Let M*_∞_ = (*s*_∞_, *r*_∞_) *be the limit at infinity. Then*

- *M*_∞_ *depends only on the energy h*_0_(*M*_0_), *i*.*e. there exists a function g* : [0, +∞[→ *T with M*_∞_ = *g*(*h*_0_(*M*_0_)).
- *r*_∞_ *is an increasing function of the energy h*_0_(*M*_0_). *The relations r*_∞_ *> r*_*herd*_ *and* ln(1 − *r*_∞_) + *R*_0_*r*_∞_ = ln(*s*(*M*_0_)) + *R*_0_*r*(*M*_0_) *characterise r*_∞_.

*Proof*. We have shown in Theorem 7 and Proposition 10 that in the case *i >* 0, the trajectory is non constant, and *M*_∞_ is characterised by 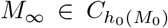 and *r*(*M*_∞_) ≥ *r*_*herd*_. This proves that *M*_∞_ depends only on *h*_0_(*M*_0_). The energy of the point *M*_∞_ = (*s*_∞_, *r*_∞_) is up to a constant − ln(1 − *r*_∞_) − *R*_0_*r*_∞_. This energy is a strictly increasing function of *r*_∞_ on the domain *r*_∞_ ∈ [*r*_*herd*_, 1[.

#### Theorem 17.

*Let β*(*t*) *be a control and M* (*t*) *a solution of the associated sir-system. Let* 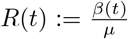 *be the propagation number induced by the mitigation and let h*_0_(*t*) *be the energy at time t. Then* 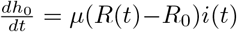.*For a mitigation at time t, the instantaneous power* 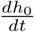 *is negative*.

*Proof*. By definition of *h*_0_(*t*), we have 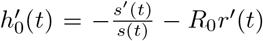. By the sir system, 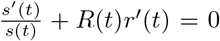 and *r*^*′*^(*t*) = *μi*(*t*). The formula for 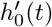 follows. A mitigation at time *t* satisfies (by definition) *R*(*t*) *< R*_0_ thus the power is negative.

### 4.6 Trajectories and solutions

Starting with a parameterised solution *t* ↦ (*s*(*t*), *i*(*t*), *r*(*t*)), ℝ → ℝ^3^ of a controlled sir-system, we can forget the time *t* keeping only the non parameterised curve *C* ⊂ ℝ^3^ which is the image of the solution. The time parametrisation is apparently lost by this reduction, but this is not true. We can recover the time *t* (up to translation) and even the control *β*(*t*) that induces the trajectory *C* (Theorem 20). This fact will be used in the next sections to compare how coercive are various strategies : We will recover *β*(*t*) from the trajectories associated to the strategies.

We work as before in the (*s, r*)-plane with *C* ⊂ ℝ^2^ instead of *C* ⊂ ℝ^3^ since *i* = 1 − *s* − *r*. At the level of the differential equation, we are interested in the system (∗∗) formulated using only the variables *r* and *s* :

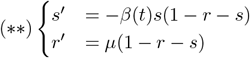

#### Definition 18.

*An infinite trajectory of the system* (∗∗) *is a set C* ⊂ ℝ^2^ *image of t* → (*s*(*t*), *r*(*t*)), *where* (*s, r*) *is a solution of* (∗∗) *defined for t* ∈ [0, +∞[*and satisfying* (*s*(0), *r*(0)) ∈ *T*.

#### Remark 19.

*The equations in* (**) *are the equations we use throughout this article. As long as a model carries explicitly or implicitly these equations, the results of the present paper apply. This is the case for the sird model with deaths which boils down to a simple sir model if the deaths and the recovered are gathered in a unique compartment. We also expect that the same approach and methods would lead to similar results as long as we can eliminate time dt from the equations to get an equation with separate variables* 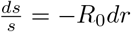 *dr leading to the same associated foliation. This the case for instance for the seir model without vital dynamics*.

#### Theorem 20.

*Let C* ⊂ *T be a subset. The following conditions are equivalent*.

1. *C is a non constant infinite trajectory of a sir-controlled system for some control function β*(*t*),
2. *• r*(*C*) *is a semi-closed interval* [*r*_0_, *r*_∞_[
  - *There exists a function* 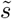: [*r*_0_, *r*_∞_[→ ℝ *continuous, piecewise C*^1^, *decreasing, satisfying* 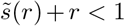, *whose graph* 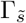 *is the set C*,
  - *The limit s*_∞_ := 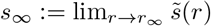 *satisfies s*_∞_ + *r*_∞_ = 1.

*Moreover, we have the formulas* 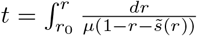 *and* 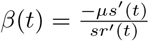.

*Proof*. We prove first that 1 ⇒ 2. By Theorem 3, a solution *s*(*t*) is decreasing and piecewise *C*^1^ (with respect to t). The sir-equation *r*^*′*^ = *μi* implies that *r* = *r*(*t*) is an increasing *C*^1^ diffeomorphism from *t* ∈ [0, ∞[onto *r* ∈ [*r*_0_, *r*_∞_[. By inversion we obtain a function *t* = *t*(*r*) which is *C*^1^ and strictly increasing. The function *s* of *t* can then be expressed with the parameter *r* letting 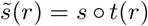. The graph and the limits stay unchanged by this reparametrisation with *r* instead of *t* and the equalities *s*_∞_ + *r*_∞_ = 1 and 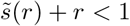 follow from the corresponding results for *s* in Theorem 3.

Conversely, we let 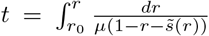. Since 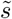 is decreasing, it follows 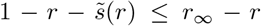 and then 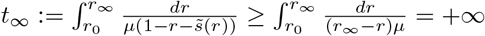. This makes *t* : [*r_0_, r*_∞_[→ [0, ∞[a strictly increasing *C* function of *r*. We denote by *r* = *r*(*t*) : [0, ∞[→ [*r*_0_, *r*_∞_[the inverse function which is *C*^1^ too. We let 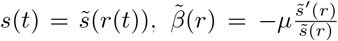, and 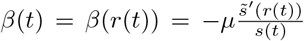. We claim that *s*(*t*), *r*(*t*) is the solution of the sir-system with control *β*(*t*) with initial condition 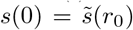 and *r*(0) = *r*_0_, hence gives a parametrisation of *C* as a trajectory. This is a direct computation: The formula for *t* shows that 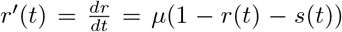 and 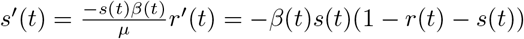.

### 4.7 Cost functions

Some people prefer long loose interventions, while others prefer short harsh interventions. Mathematically, we encode the preferences with cost functions. The cost increases when mitigations become more intensive or when the duration increases. Each person carries its own cost function.

#### Definition 21.

*Recall that a sir model comes with a constant β*_0_ *depending on biological conditions. Let β be a control with values in* [0, *β*_0_]. *The number β*(*t*) *is called the constraint at time t. There is no constraint if β*(*t*) = *β*_0_ *and there is a constraint if β*(*t*) *< β*_0_. *The maximal constraint for the control β is the infimum infess*(*β*(*t*)), *where infess denotes the essential infimum*.

*A cost function is a decreasing function c* : [0, *β*_0_] → ℝ *with c*(*β*_0_) = 0.

*The duration d*(*β*) *of a control is the Lebesgue measure of the locus β*^−1^([0, *β*_0_[).

*The cost of a control β is* 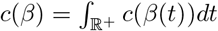. *A time interval I such that there is no constraint outside I is called a constraint interval. The cost function can be computed after restriction to a constraint interval* : *For any constraint interval I, i*.*e. c*(*β*) = ∫_*I*_ *c*(*β*(*t*)).

*A control β*_1_ *is preferable to a control β*_2_ *(In notation* : *β*_1_ ≥ *β*_2_*) if r*_∞_(*β*_1_) ≤ *r*_∞_(*β*_2_) *and for every cost function c, c*(*β*_1_) ≤ *c*(*β*_2_). *The relation* ≤ *is a partial preorder. A control β*_1_ ∈ 𝒞 *is optimal in the set of controls* 𝒞 *if for every β*_2_ ∈ 𝒞, *we have β*_1_ ≥ *β*_2_. *A control β*_1_ ∈ 𝒞 *is maximal in C if there is no better control in* 𝒞 : *for all β*_2_ ∈ 𝒞 *we have β*_1_ ≥ *β*_2_ *or β*_1_ *and β*_2_ *incomparable*.

#### Remark 22

*With informal words, an optimal control is a control preferred by everyone regardless of the subjective personal cost function, with r*_∞_ *minimal. A maximal control is a control β which cannot be improved: replacing β by β*^*′*^ *increases r*_∞_ *or makes at least one person dissatisfied by a higher cost*.

*If there is an optimal control in a set* 𝒞 *of considered choices, the public policy is obviously chosen* : *an optimal control is a universal favourite choice for all people. If no optimal control exists, then there is no universal choice and political arbitrations are required. However, there are still choices that can be improved universally* : *If everyone prefers β*_2_ *than β*_1_, *the public policy should reject β*_1_, *in favour of β*_2_. *At the end, the public choice should be ideally between controls that can’t be universally ameliorated any more, i*.*e. an option is selected between the maximal controls*.

#### Remark 23.

*We have used the essential infimum rather than the infimum in the definition of the maximal constraint because of the points a*_*i*_ *where β may be discontinuous*.

*We underline that the order for controls is different from the order for functions. The relation β*_1_ ≤ *β*_2_ *as controls defined above does not imply that β*_1_ ≤ *β*_2_ *as usual functions*.

The following theorem gives necessary conditions to compare two controls. If *β*_2_ is universally preferred to *β*_1_, then the mitigation is shorter-lived and the maximal constraint is smaller.

#### Theorem 24.

*Let 𝒞 be a set of controls with the same r*_∞_ : ∀*β*_1_, *β*_2_ ∈ *C, r*_∞_(*β*_1_) = *r*_∞_(*β*_2_). *Let β*_1_, *β*_2_ ∈ *C. If β*_1_ ≤ *β*_2_, *then the duration of constraint satisfy d*(*β*_1_) ≥ *d*(*β*_2_), *and the maximum of constraints satisfy infess*(*β*_2_(*t*))) ≥ *infess*(*β*_1_(*t*)).

*Proof*. Consider the cost function *c*(*x*) which is equal to 1 if *x* ∈ [0, *β*_0_[and 0 otherwise; then *c*(*β*) = *d*(*β*). If *β*_2_ ≥ *β*_1_, *d*(*β*_1_) = *c*(*β*_1_) ≥ *c*(*β*_2_) = *d*(*β*_2_).

For the second inequality, we suppose ad absurdum that *infessβ*_1_ *> infessβ*_2_ and we take *β*_3_ with *infessβ*_1_ *> β*_3_ *> infessβ*_2_. Then we choose the cost function which is 1 in [0, *β*_3_] and 0 otherwise. Then *c*(*β*_1_) = 0 *< c*(*β*_2_).

Intuitively, if we have two controls *β*_1_ and *β*_2_, and if the constraints in *β*_2_ are harsher and longer than those for *β*_1_, then the cost functions should satisfy *c*(*β*_2_) ≥ *c*(*β*_1_). This idea is formalised with a suitable diffeomorphism in the next theorem.

#### Theorem 25.

*Let β*_1_ *and β*_2_ *be two controls and I*_1_, *I*_2_ *two constraint intervals for β*_1_ *and β*_2_. *Suppose that there exists a diffeomorphism φ* : *I*_1_ → *I*_2_ *with φ*^*′*^(*t*) ≥ 1 *and β*_2_(*φ*(*t*)) ≤ *β*_1_(*t*). *Then for any cost function c, c*(*β*_2_) ≥ *c*(*β*_1_). *In particular, if r*_∞_(*β*_2_) ≤ *r*_∞_(*β*_1_), *then β*_2_ ≤ *β*_1_.

*Proof*. We have 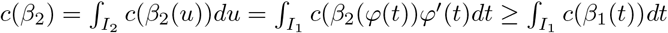.

### 4.8 Strategies with constant mitigation

In this section, we consider constant mitigations, that is mitigations with a fixed level of intervention and we investigate the problem of their optimality : which ones are optimal, which ones are not ?

A first approach with constant mitigations is to minimise the burden for a fixed duration. For instance, we may consider two strategies for promoting remote work : The first strategy promotes remote work every Monday for five weeks. The second strategy promotes remote work from Monday to Friday after one month.

Both strategies have the same type of restriction, and the same duration (5 days). Which strategy yields to the smallest ratio *r*_∞_ of infected people ? More generally, keeping the same interventions and the same duration, and changing the scheduling, is there a mitigation strategy minimising the burden of the epidemic?

An other approach with constant mitigations could be to fix a maximal burden *r*_∞_ and a level of mitigation (in the example above, remote work). Then the policy makers try to minimise the mitigation time required to keep *r*_∞_ under the chosen limit. What are the optimal mitigations for this question ?

We will prove in this section that the two above problems (minimising the duration for a fixed burden or minimising the burden for a fixed duration) are equivalent : a strategy based on a constant control *β* is optimal for one problem if and only if it is optimal for the other problem (Theorem 28). Such an optimal *β* always exists and it has specific properties. More precisely, we show that for an optimal strategy, all the mitigations must be grouped in a unique intervention. In a sir-controlled model, several short interventions are less efficient than a long adequately planned intervention of the same total duration. The adequate planning boils down to “as soon as possible” if the herd ratio has been passed, and around the zone *s* = *s*_*herd*_ otherwise.

Our argument consists in replacing the infinite dimensional space in which *β* moves with a compact space. This reduction relies on the comparison of the time spent on parallel *R*-leaves of a quadrilateral. Once the ambient space is compact, some continuity argument can be applied.

Technically, a constant mitigation appears as a control *β* that takes two values *β*_0_ and *β*_1_, where *β*_0_ is the value of *β* when no mitigation occurs, and *β*_1_ *< β*_0_ is the value of *β* during the mitigation.

#### Definition 26.

*Let β*_1_ ∈ [0, *β*_0_[. *A control β of order k* ≥ −1 *is a piecewise constant function β* : [0, +∞[→ {*β*_0_, *β*_1_} *such that there exists elements* (*a*_*i*_, *b*_*i*_)_*i*∈{0…*k*}_ *with* 0 ≤ *a*_0_ *< b*_0_ *< a*_1_ *< b*_1_ *< · · · < a*_*k*_ *< b*_*k*_, *β*(*t*) = *β*_1_ *on any interval* [*a*_*i*_, *b*_*i*_] *and β*(*t*) = *β*_0_ *otherwise. A mitigation with a control of some order k is called a constant mitigation. Its duration is d*(*β*) = Σ_*i*_(*b*_*i*_ − *a*_*i*_). *By convention, for k* = − 1, *we have β*(*t*) = *β*_0_ *for every t and d*(*β*) = 0.

*In this section, we fix once and for all an initial point M*_0_ ∈ *T, with i*(*M*_0_) *≠* 0, *and we denote by r*_∞_(*β*) *the share of people finally removed for the sir-control system with initial condition M*_0_ *and control β*.

*Recall that H*_0_(*s, r*) = ln(*s*) + *R*_0_*r with* 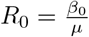. *Similarly, we let H*_1_(*s, r*) = ln(*s*) + *R*_1_*r, with* 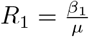.

We will consider minimisation problems where an optimal *β* ∈ 𝒞 has to be found within various classes *C*. These classes *C* are introduced in the following definition.

#### Definition 27.

*Let d* ≥ 0, *r*_∞_ ∈]*r*_*herd*_, 1[.

- 𝒞_*d,k*_ *is the set of controls β with constant mitigation of order k and duration d*(*β*) = *d*,
- 𝒞_*d*_ := ∪_*k*≥−1_𝒞_*d,k*_ *(note that* 𝒞_0_ = 𝒞_0,−1_ *and for d >* 0, 𝒞_*d*_ = ∪_*k*≥0_𝒞_*d,k*_ *since* 𝒞_*d*,−1_ *is empty)*
- 𝒞 := ∪_*d*≥0_𝒞_*d*_ *is the set of finite time constant controls*,
- 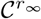 *is the set of β* ∈ 𝒞 *with r*_∞_(*β*) = *r*_∞_,
- 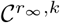 *is the set of β* ∈ 𝒞 *of order k with r*_∞_(*β*) = *r*_∞_
- 𝒞_*cross*_ *is the set of β* ∈ 𝒞 *such that there exists i, there exists t* ∈ [*a*_*i*_, *b*_*i*_] *with s*(*t*) = *s*_*herd*_. *In non technical terms, there is an alternation of mitigated and non mitigated periods, and the herd ratio s*_*herd*_ *is crossed during a mitigated period* [*a*_*i*_, *b*_*i*_] *rather than during a non mitigated period*]*b*_*i*_, *a*_*i*+1_[.
- 𝒞_*imm*_ *is the set of controls β* ∈ 𝒞 *such that a*_0_ = 0. *The exponent “imm” stands for immediate, to denote the controls that start the mitigation at t* = 0.
- *Several exponents correspond to intersections of the classes above. For instance*, 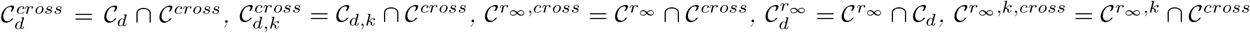.

#### Theorem 28.

1. *Let β* ∈ 𝒞, *d* = *d*(*β*), *and r*_∞_ = *r*_∞_(*β*). *Then β is optimal in* 𝒞_*d*_ *if and only if β is optimal in* 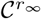.
2. *For every d* ≥ 0, *there exists β* ∈ 𝒞_*d*_ *which is optimal in* 𝒞_*d*_.
3. *For every r*_∞_ *such that* 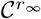 *is non empty, there exists β* ∈ 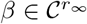 *which is optimal in* 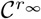.
4. 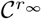 *is non empty if and only if r*_∞_ ∈]*r*_∞_(*β*_1_), *r*_∞_(*β*_0_)] *where*
  - *r*_∞_(*β*_0_) *is the limit of r*(*t*) *when β*(*t*) = *β*_0_ *for all t (no mitigation)*.
  - *r*_∞_(*β*_1_) = lim_*N*→∞_ *r*_∞_(*β*_1,*N*_) *where β*_1,*N*_ (*t*) = *β*_1_ *for all t* ≤ *N, and β*_1,*N*_ (*t*) = *β*_0_ *for t > N*. *In intuitive terms, r*_∞_(*β*_1_) *is the ratio of infected people after an arbitrarily long but still finite mitigation whose intensity is governed by β*_1_.
5. *We denote by r*_∞_(*d*) *the quantity r*_∞_(*β*) *when β is optimal in C*_*d*_ *and by d*(*r*_∞_) *the quantity d*(*β*^*′*^) *where β*^*′*^ *is optimal in* 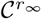. *Then the functions d* → *r*_∞_(*d*) *and r*_∞_ → *d*(*r*_∞_) *are mutually inverse for d* ∈ [0, +∞[*and r*_∞_ ∈]*r*_∞_(*β*_1_), *r*_∞_(*β*_0_)].
6. *If β* ∈ 𝒞 _*d*_ *is optimal, then β* ∈ 𝒞_*d*,0_. *In other words, an optimal mitigation does not split the mitigation in parts*.
7. *If β is optimal and s*(*M*_0_) ≤ *s*_*herd*_, *then β* ∈ 𝒞_*imm*_. *In other words, the optimal mitigation starts as soon as possible if s*(*M*_0_) ≤ *s*_*herd*_.
8. *If β is optimal and s*(*M*_0_) ≥ *s*_*herd*_, *then β* ∈ 𝒞_*cross*_. *In other words, the adequate scheduling of the mitigation is such that the vertical line s* = *s*_*herd*_ *is crossed during the mitigation* : *the mitigation occurs around the zone where s* = *s*_*herd*_.

We start the proof with a lightened form of the theorem, considering only one step mitigations (*β* of order 0) and *s*(*M*_0_) ≥ *s*_*herd*_.

#### Proposition 29.

*We suppose s*(*M*_0_) ≥ *s*_*herd*_.

1. *Let* 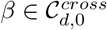, *and r*_∞_ = *r*_∞_(*β*). *Then β is optimal in* 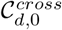 *if and only if β is optimal in* 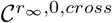.
2. *For every d* ≥ 0, *there exists* 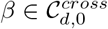 *which is optimal in* 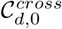.
3. *For every r*_∞_ *such that* 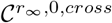 *is non empty, there exists an optimal* 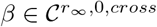 *which is optimal in* 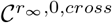.
4. 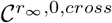 *is non empty if and only if r*_∞_ ∈]*r*_∞_(*β*_1_), *r*_∞_(*β*_0_)] *where*
  - *r*_∞_(*β*_0_) *is the limit of r*(*t*) *when β*(*t*) = *β*_0_ *for all t (no mitigation)*.
  - *r*_∞_(*β*_1_) = lim_*N*→∞_ *r*_∞_(*β*_1,*N*_) *where β*_1,*N*_ (*t*) = *β*_1_ *for all t* ≤ *N, and β*_1,*N*_ (*t*) = *β*_0_ *for t > N*.
5. *Denote by r*_∞_(*d*) *the quantity r*_∞_(*β*) *when β is optimal in* 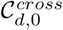 *and denote by d*(*r*_∞_) *the quantity d*(*β*^*′*^) *where β*^*′*^ *is optimal in* 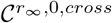. *Then the functions d* → *r*_∞_(*d*) *and r*_∞_ → *d*(*r*_∞_) *are mutually inverse for d* ∈ [0, +∞[*and r*_∞_ ∈]*r*_∞_(*β*_1_), *r*_∞_(*β*_0_)].

*Proof*. From the definition of optimality, we have *β* optimal in 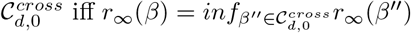. Indeed, all *β* ∈ *C*_*d*_ have the same cost *c*(*β*_1_)*d* for every cost function *c*. Thus *β < β*^*′*^ as controls if and only iff *r*_∞_(*β*) *> r*_∞_(*β*^*′*^). Similarly, *β* optimal in 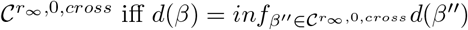.

Suppose that *β* is not optimal in 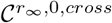, then there exists *β*^*′*^ with *d*(*β*^*′*^) *< d*(*β*) and *r*_∞_(*β*^*′*^) = *r*_∞_.

The control *β*^*′*^ has value *β*_1_ on an interval [*a*_0_, *b*_0_] with *b*_0_ *< a*_0_ + *d*(*β*). Define *β*^*′′*^ by *β*^*′′*^(*t*) = *β*_1_ for *t* ∈ [*a*_0_, *a*_0_ + *d*(*β*)] and *β*^*′′*^(*t*) = *β*_0_ otherwise. Since there is a longer mitigation in *β*^*′′*^ than in *β*^*′*^, it follows that *r*_∞_(*β*^*′′*^) *< r*_∞_(*β*^*′*^) = *r*_∞_. Thus *β* is not optimal in 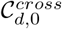 since *β*^*′′*^ is a better choice.

Suppose conversely that *β* is not optimal in 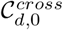. Then there exists *β*^*′*^ with *r*_∞_(*β*^*′*^) *< r*_∞_(*β*) and *d*(*β*^*′*^) = *d*. The control *β*^*′*^ has value *β*_1_ on an interval [*a*_0_, *a*_0_ + *d*]. Define 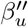 depending on *u* ∈ [0, *d*] by 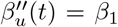 for *t* ∈ [*a*_0_, *a*_0_ + *u*] and *β*^*′′*^(*t*) = *β*_0_ otherwise. For *u* = 0, there is no mitigation thus 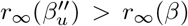. For *u* = *d*, 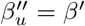, thus the opposite inequality holds. By the intermediate value theorem, there exists *u*_0_ ∈]0, *d*[with 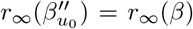. If 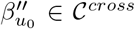, then *β* is not optimal in 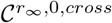 since 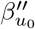 is a better choice. If 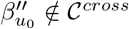, then we replace 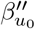 by *β*^*′′′*^ ∈ 𝒞_*cross*_ using proposition 35. This concludes the proof of item 1.

Now we prove item 2. Let *β*_*no*_ = *β*_0_ be the constant control corresponding to no mitigation and let *t*_*herd*_ be such that *s*(*t*_*herd*_) = *s*_*herd*_ for the sir-system defined by *β*_*no*_. The existence of *t*_*herd*_ follows from the hypothesis *s*(*M*_0_) ≥ *s*_*herd*_ and from the equality *s*_∞_ *< s*_*herd*_ (Theorem 16). For *u* ∈ [0, *t*_*herd*_], define *β*_*u*_ by *β*_*u*_(*t*) = *β*_1_ for *t* ∈ [*u, u* + *d*], and *β*_*u*_(*t*) = *β*_0_ otherwise. Let *s*_*u*_(*t*) be the function *s* associated to the sir-system with control *β*_*u*_. Then *β*_*u*_ ∈ *C*^*cross*^ if and only if *s*_*u*_(*u* + *d*) ≤ *s*_*herd*_. By continuity in *u*, this is a closed condition on the parameter *u* ∈ [0, *t*_*herd*_], thus *u* lives in a compact *K*. It follows that the elements 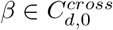 are parameterised by an element *u* ∈ *K*. The map *K* → ℝ, *u* ↦ *r*_∞_(*β*_*u*_) is a continuous function on *K*, hence it admits a minimum. This proves item 2.

Item 4 is easy.

For every *d* ∈ *D* = [0, +∞[, the set 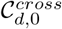 contains an optimal control *β* by item 2. Let *B* ⊂ [0, 1] be the set of elements *r*_∞_ such that 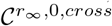 contains an optimal control. Let *β* ∈ 𝒞 be a control of order 0.

If *β* is optimal (equivalently in 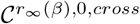 or 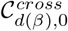), then *d*(*β*) ∈ *D, r*_∞_(*β*) ∈ *B*, and *d*(*r*_∞_(*β*)) = *d*(*β*) and *r*_∞_(*d*(*β*)) = *r*_∞_(*β*). This proves that the functions *r*_∞_ and *d* in item 5) are mutually inverse one-to-one correspondences between *D* = [0, +∞[and *B*. Since *r*_∞_(*d*) is a continuous strictly decreasing function of *d*, we have *B* =] lim_*d*→∞_ *r*_∞_(*d*), *r*_∞_(0)]. A mitigation of duration 0 means no mitigation, thus *r*_∞_(0) = *r*_∞_(*β*_0_). The equality lim_*d*→∞_ *r*_∞_(*d*) = *r*_∞_(*β*_1_) holds by definition of *r*_∞_(*β*_1_) = lim_*N*→∞_ *r*_∞_(*β*_1,*N*_) since *r*_∞_(*N*) ≤ *r*_∞_(*β*_1,*N*_) and *r*_∞_(*d*) ≥ *r*_∞_(*β*_1,*N*_) for *N* large enough. This concludes the proof of item 5 and item 3.

To prove Theorem 28, the main point now is to show that we can reduce to the simple version we proved in Proposition 29. That is, we need to show that an optimal control *β* corresponds to a one step mitigation and is in 𝒞^*cross*^. This will be done in Proposition 35. Proposition 35 requires some lemmas relative to *R*_0_ − *R*_1_ quadrilaterals that we define and study now.

#### Definition 30.

*A R*_0_ − *R*_1_ *quadrilateral is a set of* 4 *points a, b, c, d* ∈ *T with*

- *a, b are on a common R*_0_*-leaf C*_*ab*_,
- *c, d are on a common R*_0_*-leaf C*_*cd*_,
- *a, d are on a common R*_1_*-leaf C*_*ad*_,
- *b, c are on a common R*_1_*-leaf C*_*bc*_,

The above definition does not fix the order of the 4 points *a, b, c, d* of the quadrilateral. The following lemma indicates how the order can be fixed.

#### Lemma 31.

*In a R*_0_ − *R*_1_ *quadrilateral, the following conditions are equivalent:*

1. *r*(*a*) = *max*{*r*(*a*), *r*(*b*), *r*(*c*), *r*(*d*)}
2. *r*(*c*) = *min*{*r*(*a*), *r*(*b*), *r*(*c*), *r*(*d*)}.
3. *H*_1_(*a*) = *H*_1_(*d*) *< H*_1_(*c*) = *H*_1_(*b*) *and H*_0_(*a*) = *H*_0_(*b*) *> H*_0_(*d*) = *H*_0_(*c*).

*Proof*. For *p* = (*s*_*p*_, *r*_*p*_), *q* = (*s*_*q*_, *r*_*q*_), we have (*H*_0_(*p*) − *H*_0_(*q*)) − (*H*_1_(*p*) − *H*_1_(*q*)) = (*R*_0_ − *R*_1_)(*r*_*p*_ − *r*_*q*_). Since *R*_0_ *> R*_1_, the equivalences 1 ⇔ 3 and 2 ⇔ 3 follow easily.

#### Lemma 32

(Completion of a triangle to a quadrilatere.). *Let a, b, c* ∈ *T with H*_0_(*a*) = *H*_0_(*b*) *> H*_0_(*c*) *and H*_1_(*b*) = *H*_1_(*c*) *> H*_1_(*a*). *Then there exists d* ∈ *T such that a, b, c, d is a R*_0_ − *R*_1_ *quadrilateral satisfying the ordering of lemma 31*.

*Proof*. By the intermediate value theorem, there exists *e* ∈ *T* with *r*(*e*) = *r*(*c*), *s*(*e*) ≤ *s*(*c*), and *H*_1_(*e*) = *H*_1_(*a*). Then *H*_0_(*e*) ≤ *H*_0_(*c*) *< H*_0_(*a*). The *R*_1_-leaf containing *a* and *e* is connected by Theorem 7. When joining *e* to *a* in the *R*_1_-leaf, we find by the intermediate value theorem a point *d* with *H*_0_(*d*) = *H*_0_(*c*).

The previous lemma recovered *d* from *a, b, c*. We have an other completion lemma which has a similar proof, which recovers *b* from *a, c, d* when *s*(*c*) *< s*_*herd*_.

#### Lemma 33

(Completion of a triangle to a quadrilatere.). *Let a, c, d* ∈ *T with H*_0_(*a*) *> H*_0_(*c*) = *H*_0_(*d*) *and H*_1_(*c*) *> H*_1_(*a*) = *H*_1_(*d*). *Suppose moreover that s*(*c*) *< s*_*herd*_. *Then there exists d* ∈ *T such that a, b, c, d is a R*_0_, *R*_1_*-quadrilateral satisfying the ordering of lemma 31*.

*Proof*. Sketch. From *c*, we draw a vertical line towards the line *i* = 0 to build a point *e*. We then conclude as in the proof of lemma 32.

The following lemma compares the duration of the mitigation on two opposite edges of a quadrilateral.

#### Lemma 34.

*Let a, b, c, d be a quadrilateral satisfying the ordering of lemma 31 and suppose that s*(*a*) ≥ *s*_*herd*_. *Consider the oriented curves*

- *C*_*ba*_, *C*_*cd*_ *the R*_0_*-curves joining a to b and c to d*
- *C*_*da*_,*C*_*cb*_ *the R*_1_*-curves joining d to a and c to b*.

*Then the time spent on C*_*cb*_ *is strictly longer than the time spent on C*_*da*_.

*If the hypothesis s*(*a*) ≥ *s*_*herd*_ *is removed and replaced with s*(*c*) ≤ *s*_*herd*_, *then the opposite conclusion holds* : *strictly less time is spent on C*_*cb*_ *than on C*_*da*_.

*Proof*. By Theorem 17, the time spent on *C*_*cb*_ is 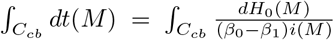. Thus, up to a positive multiplicative constant, the time is measured by integrating the differential form 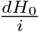. Denote by *φ* : *C*_*cb*_ → *C*_*da*_ the diffeomorphism which sends the point *M* of *C*_*cb*_ to the point *M*^*′*^ of *C*_*da*_ with *H*_0_(*M*) = *H*_0_(*M*^*′*^). By construction, *H*_0_ = *H*_0_ *° φ*. This implies *dH*_0_ = *φ*^∗^(*dH*_0_). Also, using *φ* as a change of variable, the time spent on *C*_*da*_ is

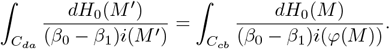

The result follows since *i*(*φ*(*M*)) is larger than *i*(*M*) by proposition 4.

#### Proposition 35.

*Let* 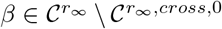 *and suppose that s*(*M*_0_) ≥ *s*_*herd*_. *Then there exists* 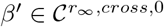 *with d*(*β*^*′*^) *< d*(*β*).

*Proof*. Suppose that *β* ∉ 𝒞^*cross*^. We want to construct *β*_1_ ∈ 𝒞^*cross*^ with the same *r*_∞_ and a smaller duration using lemma 34. The trajectory of the solution associated to *β* is characterised by a sequence of points *M*_0_ = *M* (0), *N*_1_ = *M* (*a*_0_), *M*_1_ = *M* (*b*_0_), *N*_2_ = *M* (*a*_1_), *M*_2_ = *M* (*b*_1_), …, *N*_*k*+1_ = *M* (*a*_*k*_), *M*_*k*+1_ = *M* (*b*_*k*_) where:

- *N*_*i*_, *M*_*i*_ are joined by a *R*_1_-leaf,
- *M*_*i*_, *N*_*i*+1_ are joined by a *R*_0_-leaf.

Since *β* ∉ 𝒞_*cross*_, *s*(*N*_1_) *< s*_*herd*_ or *s*(*M*_1_) *> s*_*herd*_. By symmetry, we consider the “right” case and we suppose *s*(*M*_1_) *> s*_*herd*_. We take *i* maximum such that *s*(*M*_*i*_) *> s*_*herd*_. There exists *t* with *s*(*t*) = *s*_*herd*_. We apply lemma 32 to *a* = *M* (*t*), *b* = *M*_*i*_, *c* = *N*_*i*_ which gives a point *d* such that *a* and *d* are on common *R*_1_-leaf, and *d* is on the *R*_0_-leaf common to *c* and *M*_*i*−1_. We consider *β*_1_ the control associated to the trajectory with characteristic points *M*_0_, *N*_1_, *M*_1_, …, *N*_*i*−1_, *M*_*i*−1_, *d, a* = *M* (*t*), *N*_*i*+1_, *M*_*i*+1_ By lemma 34, we have *d*(*β*_1_) *< d*(*β*).

Replacing *β* by *β*_1_, we may suppose that *β* ∈ 𝒞_*cross*_. If *k* = 0, we are done so we suppose *k >* 0 and we will construct *β*_2_ ∈ 𝒞_*cross*_ with *r*_∞_(*β*_2_) = *r*_∞_(*β*), *d*(*β*_2_) *< d*(*β*) and *β*_2_ has a smaller *k* than *β*. We take again the initial notations where *β* is characterised by the points *M*_0_, *N*_1_,, *N*_*k*+1_, *M*_*k*+1_. Since *β* ∈ 𝒞_*cross*_, there exists *i* ≥ 1 with *s*(*N*_*i*_) ≥ *s*_*herd*_ and *s*(*M*_*i*_) ≤ *s*_*herd*_. Since *k* + 1 ≠ 1, we have *i* 1 or *i ≠ k* + 1. By symmetry, we suppose *i >* 1. We apply lemma 32 to *a* = *N*_*i*_, *b* = *M*_*i*−1_, *c* = *N*_*i*−1_, which gives a point *d*. The point *d* is on the *R*_1_-leaf containing *N*_*i*_,*M*_*i*_, and on the *R*_0_-leaf common to *N*_*i*−1_ and *M*_*i*−2_. We consider *β*_2_ the control associated to the trajectory with characteristic points *M*_0_, *N*_1_, *M*_1_, …, *N*_*i*−2_, *M*_*i*−2_, *d, M*_*i*_, *N*_*i*+1_, *M*_*i*+1_…. By lemma 34, we have *d*(*β*_2_) *< d*(*β*).

#### Proposition 36.

*Let* 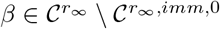 *and suppose that s*(*M*_0_) *< s*_*herd*_. *Then there exists* 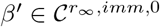 *with d*(*β*^*′*^) *< d*(*β*).

*Proof*. Similar to the proof of proposition 35.

We are now ready to prove Theorem 28.

*Proof*. Item 1) is proved as in proposition 29.

Items 6,7,8 are direct consequences of proposition 35 and 36.

For item 3, if *s*(*M*_0_) ≥ *s*_*herd*_, it follows from proposition 35 and proposition 29 that 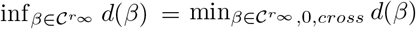. If *s*(*M*_0_) *< s*_*herd*_, proposition 36 shows that 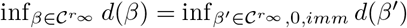. But there is a unique 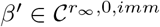, so the infimum is a minimum.

Item 4 is easy.

Let *D* ⊂ [0, +∞[be the set of elements *d* such that 𝒞_*d*_ contains an optimal control *β*. Let *B* ⊂ [0, 1] be the set of elements *r*_∞_ such that 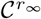 an optimal control. By item 3 and 4, we have *B* =]*r*_∞_(*β*_1_), *r*_∞_(*β*_0_)]. Let *β* ∈ *C*. If *β* is optimal (equivalently in 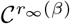 or in 𝒞_*d*(*β*)_), then *d*(*β*) ∈ *D, r*_∞_(*β*) ∈ *B*, and *d*(*r*_∞_(*β*)) = *d*(*β*) and *r*_∞_(*d*(*β*)) = *r*_∞_(*β*). This proves that the functions *r*_∞_ and *d* in item 5) are mutually inverse one-to-one correspondences. If *s*(*M*_0_) ≥ *s*_*herd*_, then 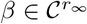 is optimal implies that 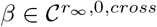. Thus *d*(*β*) has already been computed in proposition 29. In particular, we have seen in proposition 29 that *d*(*B*) = [0, +∞[. This proves item 5 and since *D* = *d*(*B*) = [0, +∞[, item 2 is proved too in the case *s*(*M*_0_) ≥ *s*_*herd*_. If *s*(*M*_0_) ≤ *s*_*herd*_, the proof is similar using the analogous of proposition 29. The analogous statement consists in replacing 𝒞_*cross*_ with 𝒞_*imm*_. The proof of the analogous proposition is basically the same. The main change is that item 2 is trivial when *s*(*M*_0_) ≤ *s*_*herd*_ since 𝒞_*imm*_ is a single point.

The following example is a confirmation of Theorem 28 in a simple case where numeric computation is possible. It considers the case of an absolute lockdown, *i*.*e. β*_1_ = 0.

#### Example 37.

*Let β*_1_ = 0. *We consider the case s*(*M*_0_) *> s*_*herd*_ *and a one step mitigation of a fixed duration d. Then the optimum control minimising r*_∞_ *is in* 𝒞^*cross*^. *In other words, the value of s is constant with s* = *s*_*herd*_ *during the mitigation*.

*Proof*. When *β*_1_ = 0 and *k* = 0, the point *M* (*t*) representing the epidemic starts at *t* = 0 on the *R*_0_-leaf defined by ln *s* + *R*_0_*r* = *c* with *c* = ln *s*(*M*_0_) + *R*_0_*r*(*M*_0_). At some time *t*, we reach a point (*s*_1_, *r*_1_) with ln *s*_1_ + *R*_0_*r*_1_ = *c* where the mitigation starts. At the end of the mitigation, solving directly the sir system, we get *M* (*t*) = (*s*_1_, *r*_2_), with *r*_2_ = *r*_1_ +*i*_1_(1 −*e*^−*μd*^) and *i*_1_ = 1−*r*_1_ −*s*_1_. To minimise *r*_∞_, we minimise the energy of *M* (*t*), or equivalently we maximise *H*(*s*_1_, *r*_2_) = ln *s*_1_ +*R*_0_*r*_2_ = ln *s*_1_ +*R*_0_*r*_1_ +*R*_0_*i*_1_(1−*e*^−*μd*^) = *c*+*R*_0_*i*_1_(1−*e*^−*μd*^). The optimum is thus obtained when *i*_1_ is maximal on the *R*_0_-curve, thus *s*_1_ = *s*_*herd*_ by proposition 4.

### 4.9 Strategies with hospital saturation

In this section, we consider situations where the health system becomes saturated when the epidemic evolves naturally from a point *M*_0_. Some mitigations are triggered to avoid the saturation that would naturally occur. Mathematically, there is a share *i*_*hosp*_ ∈ [0, 1] of infected people corresponding to a completely full but not overloaded health system. Without mitigations, the maximum *i*_*max*_ of *i*(*t*) would satisfy *i*_*max*_ *> i*_*hosp*_ and the system would be overloaded. To avoid saturation, a mitigation is triggered when some level *i*(*t*) = *i*_*trig*_ is reached, with *i*_*trig*_ ≤ *i*_*hosp*_. The ratios *i*_*hosp*_ and *i*_*max*_ are given. The ratio *i*_*trig*_ is chosen and this section discusses the choice of *i*_*trig*_ to get an efficient strategy.

In the context of monitoring the charge of the health system, some people propose to react sooner, while others propose to wait longer before launching a mitigation. The first choice corresponds to *i*_*trig*_ *<< i*_*hosp*_ and the second choice to *i*_*trig*_ = *i*_*hosp*_ − *ϵ* with *ϵ* ≥ 0 a small number. Is it preferable to react sooner or to wait ? What is the best value for *i*_*trig*_ ?

We consider two scenarios, the first one without rebound, the second one allowing a rebound of the number of infected people when the mitigation stops. In both scenarios, we suppose that at *t* = 0, the triggered level is not passed (*i*(*M*_0_) ≤ *i*_*trig*_), and that after some time, the saturation would occur in the absence of mitigation (*i*_*max*_ *> i*_*hosp*_). This implies in particular that *s*(*M*_0_) *> s*_*herd*_ (since *i* is a decreasing function of time if *s* ≤ *s*_*herd*_) and *i*(*M*_0_) *>* 0. We will assume that all these assumptions hold in this section. In summary 0 *< i*(*M*_0_) ≤ *i*_*trig*_ ≤ *i*_*hosp*_ *< i*_*max*_, and *s*(*M*_0_) *> s*_*herd*_. We will see later that we can replace the condition *i*_*trig*_ ∈ [*i*(*M*_0_), *i*_*hosp*_] with *i*_*trig*_ ∈]0, *i*_*hosp*_] when *H*(*M*_0_) ≥ 0 with a suitable change of *M*_0_ (Remark 39).

#### 4.9.1 Scenario without rebound

##### Definition 38.

*The scenario without rebound starts at t* = 0 *at a point M*_0_ *with i*(*M*_0_) *>* 0 *and s*(*M*_0_) *> s*_*herd*_. *It is defined as follows*.

- *The scenario depends on i*_*trig*_ ∈ [*i*(*M*_0_), *i*_*hosp*_] *and we denote by* 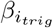 *the corresponding control*.
- *The scenario is divided in three stages t* ∈ [0, *t*_*trig*_], *t* ∈ [*t*_*trig*_, *t*_*relax*_], *t* ≥ *t*_*relax*_.
- *For t* ≤ *t*_*trig*_, *the disease evolves with no constraint:* (*β*_*trig*_(*t*) = *β*_0_) *and i*(*t*) ≤ *i*_*trig*_.
- *For t* ∈ [*t*_*trig*_, *t*_*relax*_], *the control β*_*trig*_(*t*) *(hence the mitigation policies) is calibrated so that i*(*t*) = *i*_*trig*_ *is constant on this period*.
- *For t* ≥ *t*_*relax*_, *β*_*trig*_(*t*) = *β*_0_ : *all constraints are removed*.
- *t*_*trig*_ *is defined as the smallest t such that i*(*t*) = *i*_*trig*_ *(the mitigation starts when the critical level is reached)*.
- *t*_*relax*_ *is characterised by s*(*t*_*relax*_) = *s*_*herd*_ *(the mitigation is removed when i*(*t*) *decreases naturally, so that i*(*t*) *will never exceed the critical level i*_*trig*_).

Since *M*_0_ can be replaced with an other point 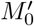 with smaller *i* without changing the problem, we can choose any *i*_*trig*_ ∈]0, *i*_*hosp*_]. Let us formalise this remark.

##### Remark 39.

*We asked for the condition i*_*trig*_ ∈ [*i*(*M*_0_), *i*_*hosp*_] *with i*(*M*_0_) *>* 0. *In fact, we can consider i*_*trig*_ ∈]0, *i*_*hosp*_] *if H*(*M*_0_) ≥ 0 *using a suitable change of M*_0_.

*Indeed, if* 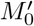 *and M*_0_ *are on the same R*_0_*-curve with* 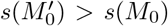 *and i*(*M*_0_) *>* 0, *then the epidemic starting at* 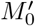 *goes through M*_0_. *It follows from the above description of the scenario that the mitigation is the same. The only difference between the two situations is that the initial part (before the mitigation occurs, in red on figure 20) is longer when the initial point is* 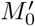. *In particular, when i*_*trig*_ *is fixed, the cost and the r*_∞_ *of the strategy is the same if we replace M*_0_ *by* 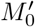.

**Figure 19:**
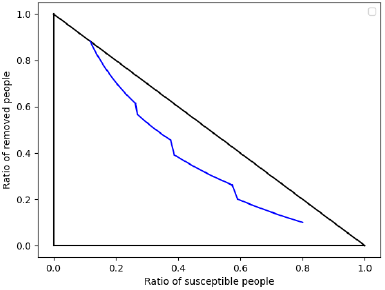
Epidemic with a control of order 2 and *M*_0_ = (0.8, 0.1).

**Figure 20:**
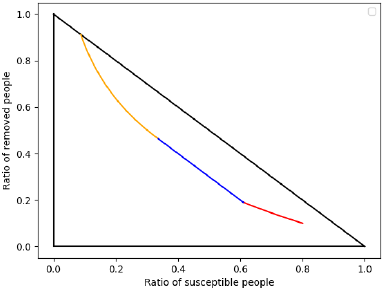
Scenario with *M*_0_ = (0.8, 0.1), *i*_*trig*_ = 0.2.

**Figure 21:**
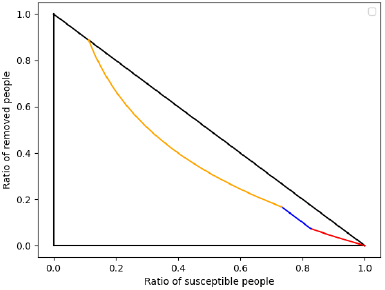
Scenario with rebound, *M*_0_ = (0.999, 0).

*As a consequence the inequality i*_*trig*_ ≥ *i*(*M*_0_) *is not necessary; the inequality* 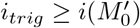 *for some* 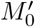 *as above is enough. If H*(*M*_0_) ≥ 0, *then i*(*M*_*init*_) = 0 *in Theorem 7, item5. Thus* 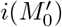 *is arbitrarily small and the required condition on i*_*trig*_ *is i*_*trig*_ *> in* 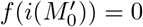.

*Since H*(1, 0) = 0 *and since mitigations increase H, the condition H*(*M*_0_) ≥ 0 *is true if M*_0_ *is a situation obtained after an epidemic has started and possibly some mitigations occurred*.

##### Theorem 40.

*Scenario without rebound*.

- *The share i*(*t*) *of infected people is increasing for t* ≤ *t*_*trig*_, *constant for t* ∈ [*t*_*trig*_, *t*_*relax*_], *decreasing for t* ≥ *t*_*relax*_.
- 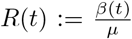 *has constant value R*(*t*) = *R*_0_ *for t* ≤ *t*_*trig*_, *is increasing continuously for t* ∈ [*t*_*trig*_, *t*_*relax*_] *from* 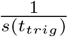 *to R*_0_, *is constant equals to R*_0_ *for t* ≥ *t*_*relax*_. *In particular, R*(*t*) *is continuous except at t* = *t*_*trig*_.
- *r*_∞_ *is a strictly increasing function of the parameter i*_*trig*_.
- *If* 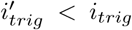, *then for every cost function c*, 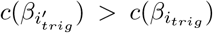. *In other words, a small i*_*trig*_ *is universally costly*.
- *Suppose that both i*_*trig*_ *and M*_0_ *are varying. The duration* 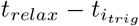 *of the mitigation tends to* +∞ *when i*_*trig*_ *tends to* 0 *and M*_0_ *stays in a region s* ≥ *s*_*min*_ *with s*_*min*_ *> s*_*herd*_.

*Proof*. First we remark that *t*_*trig*_ is well defined. Indeed, by hypothesis *i*_*max*_ *> i*_*hosp*_ ≥ *i*_*trig*_ ≥ *i*(*M*_0_) ≥ 0. Thus, without mitigation *i*(*t*) would increase from *i*(*M*_0_) to *i*_*max*_ and there is some *t* with *i*(*t*) = *i*_*trig*_ by intermediate value theorem.

In the absence of mitigation, *i*_*max*_ is realised at a point (*s, r*) with *s* = *s*_*herd*_ and *s* is a strictly decreasing function of *t*. Since the mitigation occurs before *i* = *i*_*max*_, we have *s*_*trig*_ *> s*_*herd*_. For *t* ≥ *t*_*trig*_, the trajectory is on the diagonal line *D* with equation *r* + *s* = 1 − *i*_*trig*_. It is geometrically clear that *D* intersects the vertical line *V* with equation *s* = *s*_*herd*_ in a point *N* ∈ *T*. Thus *t*_*relax*_ is well defined : it is defined by *t*_*relax*_ − *t*_*trig*_ is the time spent on the segment [*M* (*t*_*trig*_)*N*].

Since on a *R*_0_-leaf, *i*(*t*) is increasing iff *s*(*t*) ≥ *s*_*herd*_ (proposition 4), the first item holds.

The second item follows from the formula for *β* in Theorem 20, with the remarks that *s*^*′*^ = − *r*^*′*^ when *i*(*t*) is constant and that 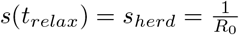.

The number *r*_∞_ is an increasing function of the energy level *h*_0_(*s, r*) associated to a *R*_0_-curve. For *t* ≥ *t*_*relax*_, *M* (*t*) lies on a fixed *R*_0_ curve, thus it suffices to compute the value of *h*_0_ at time *t* = *t*_*relax*_ to characterise *r*_∞_. Now *M* (*t*_*relax*_) = (*s*_*herd*_, *r*_*relax*_ = 1 − *s*_*herd*_ − *i*_*relax*_ = 1 − *s*_*herd*_ − *i*_*trig*_). Since the first variable *s* = *s*_*herd*_ is fixed, the formula for *h*_0_ yields that *h*_0_ is a monotonous function of *r*_*relax*_, hence of *i*_*trig*_. This proves the third.

As for the fifth item, since 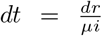 by the sir equations, we have 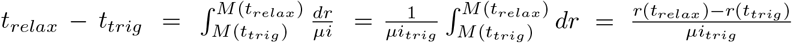. When *i*_*trig*_ (*n*) is a sequence that tends to 0, and *M*_0_ (*n*) is a sequence of initial points that stays in the zone [*s*_*min*_, 1[, then the limit (as a function of *n*) of *r*(*t*_*relax*_(*n*)) = 1−*s*(*t*_*relax*_(*n*))−*i*_*trig*_(*n*) = 1−*s*_*herd*_−*i*_*trig*_(*n*) is 1−*s*_*herd*_. Since *i*_*trig*_(*n*) tends to 0, and since in the zone *s > s*_*min*_, a *R*_0_-leaf has a tangent with slope 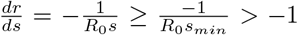, the distance between *M*_0_(*n*) and *M* (*t*_*trig*_(*n*)) tends to 0. In particular, lim sup *r*(*t*_*trig*_(*n*)) = lim sup *r*(*M*_0_(*n*)) = 1 − lim inf *s*(*M*_0_(*n*)) ≤ 1 − *s*_*min*_ *<* 1 − *s*_*herd*_. The formula for *t*_*relax*_ − *t*_*trig*_ then implies that lim_*n*→+∞_ *t*_*relax*_(*n*) − *t*_*trig*_(*n*) = +∞.

The trajectory triggering at the level *i*_*trig*_ is constrained on the segment [*M* (*t*_*trig*_)), *M* (*t*_*relax*_)]. This segment is included in a line *L* : *i* = *cte*. The same remark for 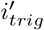 corresponds to the segment 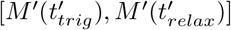 and to a line *L*^*′*^. We consider 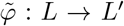 the geometrical affine map which sends *M* (*t*_*trig*_) to 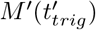 and *M* (*t*_*relax*_) to 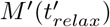. We consider *φ* the induced temporal map 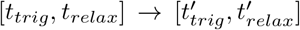 defined by 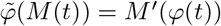. For simplicity, we use the notations *t*^*′*^ = *φ*(*t*), 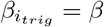 and 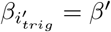. By theorem 25, to settle the fourth item, we need to prove that *β*^*′*^(*t*^*′*^) ≤ *β*(*t*) and that 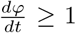. We have 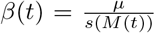 and 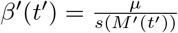 by Theorem 20. The inequality *s*(*M*^*′*^(*t*^*′*^)) *> s*(*M* (*t*)) is clear geometrically and easy to prove. The map *φ* sends [*t, t* + *dt*] to 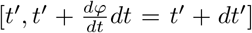. We want *dt*^*′*^ *> dt*. By the sir equations, 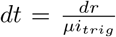 and 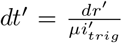. Since 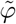 is an affine dilatation, we have *dr*^*′*^ *> dr*. Finally since 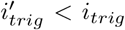, it follows that *dt*^*′*^ *> dt*.

##### Corollary 41.

*For all i*_*trig*_, *j*_*trig*_ ∈]0, *i*_*hosp*_[, *the corresponding controls* 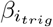 *and* 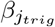 *are not comparable, thus no strategy is preferable*.

*Proof*. By Theorem 40, if *i*_*trig*_ *< j*_*trig*_, *r*_∞_ is lower for *i*_*trig*_ but at the price of a more costly mitigation.

#### 4.9.2 Scenario with rebound

The only difference between the scenario with rebound and the scenario without rebound is on the choice of *t*_*relax*_. In the previous scenario without rebound, the end of the mitigation was late, *t*_*relax*_ was chosen large so that *i*(*t*) decreases for *t* ≥ *t*_*relax*_. In the scenario with rebound, the mitigation is relaxed sooner. As a consequence, a rebound of *i*(*t*) occurs when the mitigation stops. However, the rebound must be small enough not to overload the health system. In technical terms, the inequality *i*(*t*) ≤ *i*_*hosp*_ must remain true for *t* ≥ *t*_*relax*_. This property implicitly defines *t*_*relax*_ : The mitigation is relaxed as soon as possible provided the health system is not overwhelmed by the rebound. By construction, there are less constraints for this scenario with rebound in comparison to the scenario without rebound, since this is the same level of constraints, but relaxed earlier. The formal definition of the strategy with rebound is given in the next definition.

##### Definition 42.

*The scenario with rebound starts at t* = 0 *at the point M*_0_ ∈ *T, with i*(*M*_0_) *>* 0, *s*(*M*_0_) *> s*_*herd*_. *It depends on the parameter i*_*trig*_ ∈ [*i*(*M*_0_), *i*_*hosp*_] *and it is defined via its control* 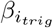 *as follows*.

- *The scenario is divided in three stages t* ∈ [0, *t*_*trig*_], *t* ∈ [*t*_*trig*_, *t*_*relax*_], *t* ≥ *t*_*relax*_.
- *For t* ≤ *t*_*trig*_, *the disease evolves with no constraint:* (*β*_*trig*_(*t*) = *β*_0_) *and i*(*t*) ≤ *i*_*trig*_.
- *t*_*trig*_ *is defined as the smallest t such that i*(*t*) = *i*_*trig*_.
- *The mitigation policies are adjusted so that i*(*t*) = *i*_*trig*_ *for t* ∈ [*t*_*trig*_, *t*_*relax*_]
- *For t* ≥ *t*_*relax*_, *β*_*trig*_(*t*) = *β*_0_ : *all constraints are removed*.
- *t*_*relax*_ *satisfies t*_*relax*_ ≥ *t*_*trig*_ *and it is the smallest such t satisfying i*([*t*, +∞[) ⊂ [0, *i*_*hosp*_] *in the absence of mitigation after t. In other words, the health system is never overwhelmed after t*_*relax*_.

Remark 39 applies here and we will can replace the condition *i*_*trig*_ ∈ [*i*(*M*_0_), *i*_*hosp*_] with *i*_*trig*_ ∈]0, *i*_*hosp*_] if *H*(*M*_0_) *>* 0.

##### Lemma 43.

*Let C*_*hosp*_ *be the R*_0_*-curve containing the point* (*s, r*) = (*s*_*herd*_, 1 − *i*_*hosp*_ − *s*_*herd*_). *Let* 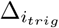 *be the line* 1 − *r* − *s* = *i*_*trig*_. *The intersection* 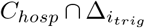 *contains a point M*_*r*_ *satisfying s*_*herd*_ *< s*(*M*_*r*_). *Moreover, M* (*t*_*relax*_) = *M*_*r*_. *In particular*, 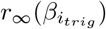 *is independent of i*_*trig*_ *as for t large enough, the point M* (*t*) *is on the R*_0_*-curve C*_*hosp*_.

*Proof*. We have parameterised any *R*_0_-leaf by a constant *c*. The *R*_0_-leaf with constant *c* has equation ln(*s*) + *R*_0_*r* = *c*. Let 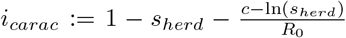. Geometrically, the meaning of *i*_*carac*_ is the following. If the *R*_0_-leaf intersects the vertical line *s* = *s*_*herd*_ in a point *M, i*_*carac*_ is the value of *i*(*M*).

Note that any *R*_0_-leaf is characterised by the value of *i*_*carac*_. The *R*_0_-leaf passing through *M* (0) = *M*_0_ has *i*_*carac*_ = *i*_*max*_ by definition of *i*_*max*_ and proposition 4. The *R*_0_-curve *C*_*hosp*_ is defined by *i*_*carac*_ = *i*_*hosp*_.

We extend the definition of *i*_*carac*_ from *R*_0_-leafs to points *M* ∈ *T* : we let *i*_*carac*_(*M*) = *i*_*carac*_(*C*) where *C* is the *R*_0_-curve through *M* (In formula : 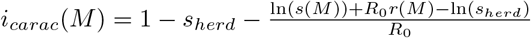.

The two points *M* (*t*_*trig*_) and *N* = (*s*_*herd*_, 1 − *s*_*herd*_ − *i*_*trig*_) are on 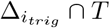. We have *i*_*carac*_(*M* (*t*_*trig*_)) = *i*_*carac*_(*M* (0)) = *i*_*max*_ and *i*_*carac*_(*N*) = *i*_*trig*_. Since *i*_*trig*_ ≤ *i*_*hosp*_ ≤ *i*_*max*_, by the intermediate value theorem, there exists *M*_*r*_ ∈ [*M* (*t*_*trig*_), *N*] with *i*_*carac*_(*M*_*r*_) = *i*_*hosp*_ (*i*.*e*.. *M*_*r*_ ∈ *C*_*hosp*_).

By definition of the strategy, the point *M* (*t*_*relax*_) is a point on the segment [*M* (*t*_*trig*_), *N*]. If *M* (*t*_*relax*_) ∈ [*M* (*t*_*trig*_), *M*_*r*_[, then *i*_*carac*_(*M* (*t*_*relax*_)) *> i*_*hosp*_ is too large and the health system is overwhelmed. If *M* (*t*_*relax*_) ∈]*M*_*r*_, *N*], then *i*_*carac*_(*M* (*t*_*relax*_)) *< i*_*hosp*_ and it is possible to reduce the duration of the mitigation with no overload on the health system, contradicting the minimality of *t*_*relax*_. Thus *M* (*t*_*relax*_) = *M*_*r*_.

##### Theorem 44.

*Scenario with rebound*.

1. *Let t*_*herd*_ *be the time such that s*(*t*_*herd*_) = *s*_*herd*_. *Then t*_*herd*_ *> t*_*relax*_. *The quantity i*(*t*) *is increasing for t* ≤ *t*_*trig*_, *constant for t* ∈ [*t*_*trig*_, *t*_*relax*_], *increasing for t* ∈ [*t*_*relax*_, *t*_*herd*_], *decreasing for t* ≥ *t*_*herd*_.
2. *R*(*t*) 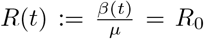 *R*_0_ *for t* ≤ *t*_*trig*_, *increasing continuously for t* ∈ [*t*_*trig*_, *t*_*relax*_] *from* 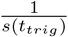 *to* 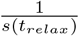, *constant equals to R*_0_ *for t* ≥ *t*_*relax*_. *In particular, R*(*t*) *is continuous except at t* = *t*_*trig*_ *and t*_*relax*_.
3. *The maximum of constraint is infess* 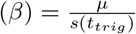. *It is an increasing function of i*_*trig*_. *Thus, from this point of view, the mitigation is harsher for a small i*_*trig*_.
4. *r*_∞_ *is constant independent of the parameter i*_*trig*_ *thus the strategies are ordered only by the cost functions*.
5. *Suppose H*(*M*_0_) ≥ 0. *There exists a unique i*_*min*_ *such that the duration* 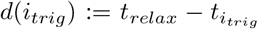 *of the mitigation as a function of i*_*trig*_ *is decreasing for i*_*trig*_ ∈]0, *i*_*min*_], *increasing for i*_*trig*_ ∈]*i*_*min*_, *i*_*hosp*_].
6. *If i*_*trig*_ *< j*_*trig*_ ≤ *i*_*min*_, *then for every cost function c*, 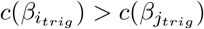. *In other words, a small i*_*trig*_ *is universally more costly*.

*Proof*. The first three items are proved like in the scenario without rebound (Theorem 40). The fourth item is a direct consequence of the previous lemma.

We now prove item 5. Since *H*(*M*_0_) ≥ 0, we suppose that *i*_*trig*_ ∈]0, *i*_*hosp*_] by remark 39.

We define *s*_1_, *s*_2_, *r*_1_, *r*_2_ functions of *i*_*trig*_ by (*s*_1_(*i*_*trig*_), *r*_1_(*i*_*trig*_)) = *M* (*t*_*trig*_) and (*s*_2_(*i*_*trig*_), *r*_2_(*i*_*trig*_)) = *M* (*t*_*relax*_).

By construction, for *k* = 1 or 2, *r*_*k*_ + *s*_*k*_ = 1 − *i*_*trig*_ and *R*_0_*r*_*k*_ + ln *s*_*k*_ = *H*_*k*_ is a constant independent of *i*_*trig*_.

By derivation with respect to *i*_*trig*_, it follows: 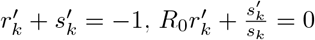, and 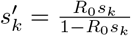.

We have 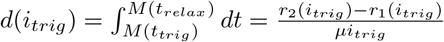, which follows from the sir-relation 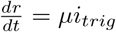. Thus *d*^*′*^ has the sign of 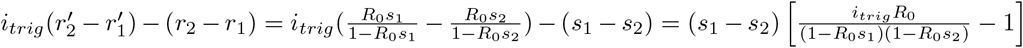. Since *s*_1_ − *s*_2_, 1 − *R*_0_*s*_1_ and 1 − *R*_0_*s*_2_ are positive, the sign of *d*^*′*^ is the sign of the function (of *i*_*trig*_) *W* = *i*_*trig*_*R*_0_ − (*R*_0_*s*_1_ − 1)(*R*_0_*s*_2_ − 1). Now we make the following remarks:

- *W* (*i*_*trig*_) is a strictly increasing function of *i*_*trig*_ ∈]0, *i*_*hosp*_]. This comes from the fact that *i*_*trig*_*R*_0_ is strictly increasing and that *s*_1_ and *s*_2_ are decreasing.
- 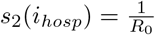 and then *W* (*i*_*hosp*_) = *i*_*hosp*_*R*_0_ *>* 0.
- The functions *s*_1_, *s*_2_ have limits *s*_1_(0) and *s*_2_(0) when *i*_*trig*_ tends to 0. We have 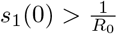 and 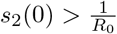. Thus the limit *W* (0) = −(*R*_0_*s*_1_(0) − 1)(*R*_0_*s*_2_(0) − 1) of *W* is strictly negative.

It follows by the intermediate value theorem that *d*^*′*^(*i*_*min*_) = 0 for some *i*_*min*_ ∈]0, *i*_*hosp*_[, and that *d*^*′*^(]0, *i*_*min*_[) ⊂] − ∞, 0[, *d*^*′*^(]*i*_*min*_, *i*_*hosp*_]) ⊂]0, +∞[. This implies the fifth item.

The last item is proved like in Theorem 40, with the following change to prove that 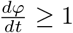, *i*.*e*. that *φ* is a local dilatation of the time. The map *φ* is affine in *r*, and since *i* is constant, the sir equation 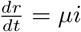 implies that *φ* is affine in *t*. It follows that *φ* is locally a dilatation of the time *t* if and only if it is globally a dilatation. Item 5 proves that *φ* is globally a dilatation and concludes the proof of item 6.

##### Corollary 45.

*For all i*_*trig*_ *< j*_*trig*_ ∈]0, *i*_*min*_], *the control* 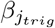 *is preferable to the control* 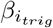. *In particular, all strategies with i*_*trig*_ *< i*_*min*_ *should be avoided*.

*For all i*_*trig*_, *j*_*trig*_ ∈ [*i*_*min*_, *i*_*hosp*_], 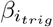 *and* 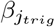 *are not comparable, thus no strategy is preferable*.

*Proof*. All the strategies with rebound have the same *r*_∞_, thus they are ordered by the cost functions. For *i*_*trig*_ *< i*_*min*_, a small *i*_*trig*_ corresponds to a universally high cost by the theorem. If *i*_*trig*_ ∈ [*i*_*min*_, *i*_*hosp*_], then a small *i*_*trig*_ corresponds to a short duration by item 5 but a high maximum constraint by item 3. Two strategies associated to *i*_*trig*_, *j*_*trig*_ ∈ [*i*_*min*_, *i*_*hosp*_] are then incomparable by Theorem 24

In summary, the theorem tells that if the mitigation occurs when the health system is insufficiently full, 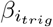 is not a maximal control and other controls are preferable. For the choice of *i*_*trig*_ to be sound, it is necessary that *i*_*trig*_ ≥ *i*_*min*_. Once *i*_*trig*_ ≥ *i*_*min*_, all choices make sense and compromises occur : a larger *i*_*trig*_ corresponds to interventions that last longer with softer maximal constraint. In particular, the control 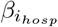 has the longest constraint time but the softest maximal constraint.

## Notes

### Competing Interest Statement

The authors have declared no competing interest.

### Funding Statement

This study did not receive any funding

